# Biological network identifies pathways linking mental health drugs to adverse weight-related side effects

**DOI:** 10.64898/2026.10.05.26364047

**Authors:** Christopher H. Arehart, Luke M. Evans

## Abstract

The majority of antidepressant and antipsychotic medications are underscored by their common weight-related adverse drug events; medication-induced weight gain is a central hindrance to psychiatric treatments because it impacts patient compliance and results in a cascade of secondary comorbidities with negative health outcomes. Here, we use network pharmacology approaches to characterize the multiple effects of drugs and explore the mechanisms by which drug-molecule interactions lead to adverse drug events. We first construct a robust interactome containing 17,293 proteins (network vertices) and 408,883 weighted interactions (network edges) and identify communities of interconnected vertices using a modularity optimization clustering algorithm. We link 33 antidepressants and 26 antipsychotics into this network via their drug-protein interactions and implement network measures of cosine similarity, shortest path distance, and connectivity significance to determine which communities are most proximal to the drugs. We identify a subnetwork of 33 (out of 536) communities enriched for genes associated with adiposity traits to test for connectivity significance to the drug vertices. Through an additional random walk analysis, we triangulate consistent links between mental health drugs and molecular pathways, including GPCR, PI3K-Akt, leptin, and insulin signaling, as well as adipogenesis. We highlight specific components within these curated pathways and map the crosstalk between them. While we demonstrate our approach in the context of mental health drugs and their weight-related side effects, our study design is broadly applicable to a wide range of drug classes, drug events, and traits of interest.

**Author Summary:** Many psychiatric medications can cause weight gain or weight loss, which can make people less likely to continue treatment and may contribute to additional health problems. Although the weight-related side effects of these medications are well known, the biological processes that connect psychiatric medications to changes in body weight are not fully understood. We developed a network-based approach to examine how psychiatric medications connect to genes associated with body weight. Our analyses identified several biological systems that consistently link psychiatric medications to weight-related biology, including systems involved in appetite regulation, insulin signaling, and the function of fat cells. More broadly, our approach provides an analytical framework for integrating information about how drugs interact with disease-associated genes/proteins to identify biological mechanisms underlying drug side effects, suggest new therapeutic targets, and support the development of safer treatments.

## Introduction

Disentangling the interconnected and abundant biological influences on human traits requires a detailed understanding of the pathways underlying them, and identifying trait-associated biomarkers can facilitate the development of drugs with targeted mechanisms of action (MOA). For instance, drugs targeting molecules with genetic evidence (e.g., significant gene-trait associations from genome wide association studies [GWASs]) are more than twice as likely to successfully progress through clinical trials.[1–4] However, it remains a challenge to characterize drugs’ many phenotypic effects and prioritize the intended (on-target) MOAs while simultaneously avoiding any unintended (off-target) drug-molecule interactions that lead to adverse drug events and potential failure during clinical trials.[5] Common approaches for connecting targets to diseases involve bipartite pairings via drug-protein interactions (DPIs);[6] however, evaluating the surrounding network of gene co-expression and protein-protein interactions (PPIs) provides important context and better captures the complex and interconnected characteristics of molecular biology.[7] In these networks (a.k.a., knowledge graphs), nodes represent functional biological units (gene products), edges represent functional associations (physical interactions or co-occurrence), and edge weights represent the confidence or strength of those associations. This network data structure is useful because it goes beyond a simple list of gene associations and models how biological components are functionally linked in a systems-level context. In addition to contextualizing PPIs and DPIs within molecular pathways, this broader network characterization can uncover aspects of genetic architecture, such as identifying and differentiating core and peripheral genes for complex traits, as suggested by the omnigenic model.[8,9] A suite of network modeling methods can be readily applied to characterize components of the network such as functional modules, mechanistic pathways, bottlenecks, central hubs, or peripheral links.

One particularly challenging example, where network-based approaches could provide insights, is the commonly reported side-effects of psychiatric medications causing weight gain and weight loss.[10] These weight-related adverse drug events (wADEs) are a central hindrance to psychiatric treatments because medication-induced weight gain impacts patient compliance with treatment plans and results in a cascade of secondary comorbidities with negative health outcomes (e.g., hypertension, dyslipidemia, type 2 diabetes, obstructive sleep apnea, and coronary artery disease).[11–15] Weight-related side effects present heterogeneously between individuals with potential mediators involving genetic effects (e.g., genetic predisposition to adiposity),[16] lifestyle factors (e.g., diet and exercise),[14] environmental variables (e.g., food availability, exposure to pollutants and chemicals),[17–20] comorbidities (e.g., eating disorders),[21] duration of therapy,[11] and rates of drug metabolism.[22] Moreover, the use of mental health drugs is widespread with 13.2% of adults in the United States being prescribed antidepressants in the past 30 days[23] and 1.6% being prescribed antipsychotics.[24] Plausibly, drug-induced wADEs involve the alteration of cellular processes orchestrating appetite regulation (e.g., leptin signaling), metabolism (e.g., insulin signaling), or sympathetic activity (e.g., sedative or stimulating effects). We hypothesized that studying drug-protein interactions in the context of an interconnected biological network context would enable the identification of key mechanisms, elucidate the biological crosstalk among relevant pathways, and highlight tissue-specific processes that are central to wADEs.

A primary advantage of studying well documented adverse side effects is the ability to infer a causal relationship between the drug’s molecular interactions and the drug event. While confounding variables can obscure some reported drug-event relationships (e.g., coinciding medications or comorbid conditions), compiled evidence supports a causal relationship when the drug precedes the event, the event resolves upon drug discontinuation, and there is a strong association with a dose-response relationship.[25] This inferred causal link between drugs and their side effects presents a unique opportunity to gain real insights into the etiology of complex polygenic diseases. When those side effects mirror a trait of interest (e.g., wADEs and adiposity), this causal link enables us to interrogate prominent gene products and implicate gene-trait associations from a more contributory angle – by building a network with links between drugs with known wADEs and genes that are associated with adiposity through prior GWAS. In addition, a minority of protein coding genes are feasibly targeted by drugs (22-26%)[26–30] and studying adiposity genetics through wADEs focuses on biological features with prior evidence for druggability, making the results directly relevant to drug development and repurposing efforts. This approach can highlight particular pathways surrounding drug targets, and even if a given drug target does not have strong evidence for genetic trait association, we can gain insights from the surrounding upstream and downstream interactions. This context provides a useful exploration of potential existing and new drug targets and generates strong hypotheses for further understanding the mechanisms by which those drugs influence traits.

In the present work, we have built upon our previous findings to further unpack the genetic architecture of adiposity and explore the mechanistic pathways linking mental health drugs to their wADEs (**Fig. 1**). In Arehart et al., 2025,[31] we disentangled the genomic architecture of adiposity and anthropometrics across the lifespan and unveiled a mostly distinct genomic component for adiposity. This genomic factor for adiposity (F4) is enriched for neuronal tissues, predicts a broad array of adverse health outcomes, and interacts with 148 drugs with wADEs. Here, we first constructed an improved biological network which excluded likely spurious interactions and rewarded redundancy (weighting interactions based on their identification and scores across multiple databases). We then prioritized communities in this network that were enriched for genes associated with adiposity and annotated them based on statistical overlap with curated pathways and biological ontology gene sets. Finally, we integrated mental health drug interactions into the network and used network measures of similarity, proximity, and topology to prioritize communities and elucidate the mechanisms through which these drugs appear to drive weight change.

**Fig. 1:**
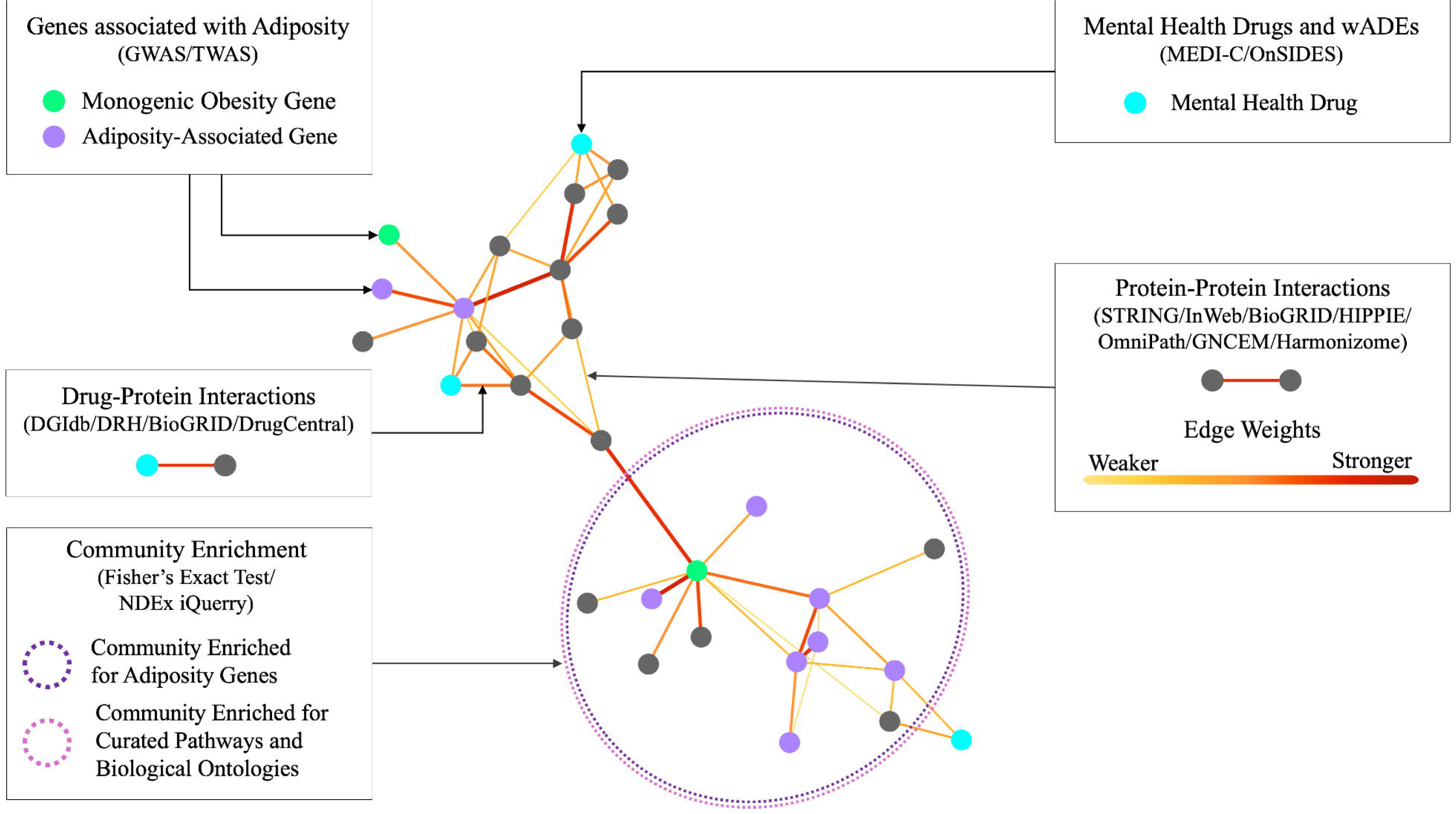
Network analysis overview. An overview of our network-based approach for identifying the mechanisms underlying the weight-related side effects of mental health drugs. This example of an annotated interactome illustrates how our improved biological network was used to model protein-protein interactions with weighted edges. Within this network we annotated vertices that corresponded to adiposity-associated genes and monogenic obesity genes. We then integrated mental health drugs through their drug-protein interactions. Through multiple analyses we identified groups of vertices that were proximal to the drugs and were enriched for adiposity-associated genes.

## Results

### Constructing a robust biological network

One challenge to leveraging biological networks for insight is that multiple such networks have been developed, but these networks only partially overlap and, when compared, exhibit notable differences mirroring their distinct methodological designs.[7,32–39] This makes it challenging to choose which network to integrate into an analysis, and it remains difficult to filter out spurious or low-confidence interactions.[40–42] An integrated, enhanced biological network is needed.

We therefore built an improved biological network using 7 databases (**Table S1, Supplementary Data 1**), primarily focusing on physical multi-validated human protein-protein interactions. Our improved network contained 408,883 interactions among 17,293 gene products, corresponding to the majority human protein coding genes. Each interaction was weighted to reflect increased evidence across different channels within and between these databases (e.g., high-throughput protein interaction assays, curated pathways, text mining, co-expression, and interlogs [**Fig. S1**]). We observed that 81% of the edges were identified by at least 2 of the 7 databases, 45% were identified by at least 3, and 14% were identified by at least 4 (**Fig. S2**). This redundancy across databases reflected a central strength of our network as we aimed to include only robust interactions and excluded nearly 5 million potential edges that were likely spurious (e.g., edges that were identified in only one database with relatively low confidence). This baseline interactome (**Fig. S3**) was agnostic relative to any phenotype, tissue, or drug interactions and modeled the broad and interconnected context of biological processes within and between cells of the human body.

### Community detection

We implemented the MONET M1 algorithm[43,44] to identify non-overlapping modules within the interactome. This clustering approach used modularity-optimized runs to identify groups of interconnected vertices for detecting community structure in our network (see Methods and **Fig. S4** for comparison to other clustering algorithms). We identified 1029 modules ranging in size from 2 to 98, and these modules contained 15,390 of the 17,293 network vertices. The median and mean module sizes were 10 and 14.96, respectively (**Fig. S5**). Many of these modules, however, were too small to meaningfully participate in downstream enrichment tests (**Fig. S6**). Thus, we designed a greedy approach to merge smaller modules with high measures of network similarity and proximity (see Methods). This ultimately resulted in 536 communities (median size = 11.50, mean size = 28.71, range = 2-199) for our downstream analyses (**Fig. 1**).

### Linking mental health drugs to the biological network

In order to identify a comprehensive set of mental health drugs, we used the MEDI-C database[45] (high precision subset) to map medications to their indications and homed in on 59 drugs indicated for mental disorders. These drugs were linked to the interactome based on 4 drug-molecule interaction databases (**Table S2; Fig. S7; Supplementary Data 2**), and interactions supported by multiple databases were given proportionally larger edge weights to reflect stronger evidence. These 59 drug vertices introduced 1710 drug-protein interactions with 289 unique protein vertices. Hereafter, we refer to the vertices predicted to interact with a drug as DrugGenes. We primarily focused on 33 antidepressants and 26 antipsychotics (**Table S3**; based on level 4 of the anatomical therapeutic chemical [ATC] classification system).

### Community proximity to mental health drugs

We implemented 2 measures of network proximity and similarity to identify neighborhoods of proteins that were closest to the mental health drugs in the network context. We used Dijkstra’s shortest distance algorithm to identify the most direct weighted path between a given drug and every protein vertex in the network (see Methods). We calculated the mean shortest distance scores of each drug to the vertices within each community. This measure of network proximity captured the topological paths between vertices but was strongly influenced by high-degree vertices that acted as connectivity hubs within the network: these connector nodes could have potentially overfit paths along spurious edges and the shortest path failed to capture the likely presence of alternative biologically plausible routes. Thus, we implemented a second measure – weighted cosine similarity – to further evaluate each vertex’s network similarity with the drugs. Cosine similarity quantified the structural equivalence between two vertices in the network using their weighted vectors from the adjacency matrix; intuitively, this cosine similarity score represented the weighted proportion of shared versus total neighbors between each protein vertex and a drug’s DrugGenes, thereby enabling us to identify vertices that had high average homophily with a drug’s targets. We then calculated the generalized mean of the shortest distance score and the cosine similarity scores for each of the 536 communities for each antipsychotic/antidepressant medication (see Methods). The two measures of proximity and similarity trended upwards together but were not entirely synonymous (**Fig. 2**. For example, some communities had relatively high mean cosine similarity scores, but modest mean shortest distance scores (and vice versa).

**Fig. 2:**
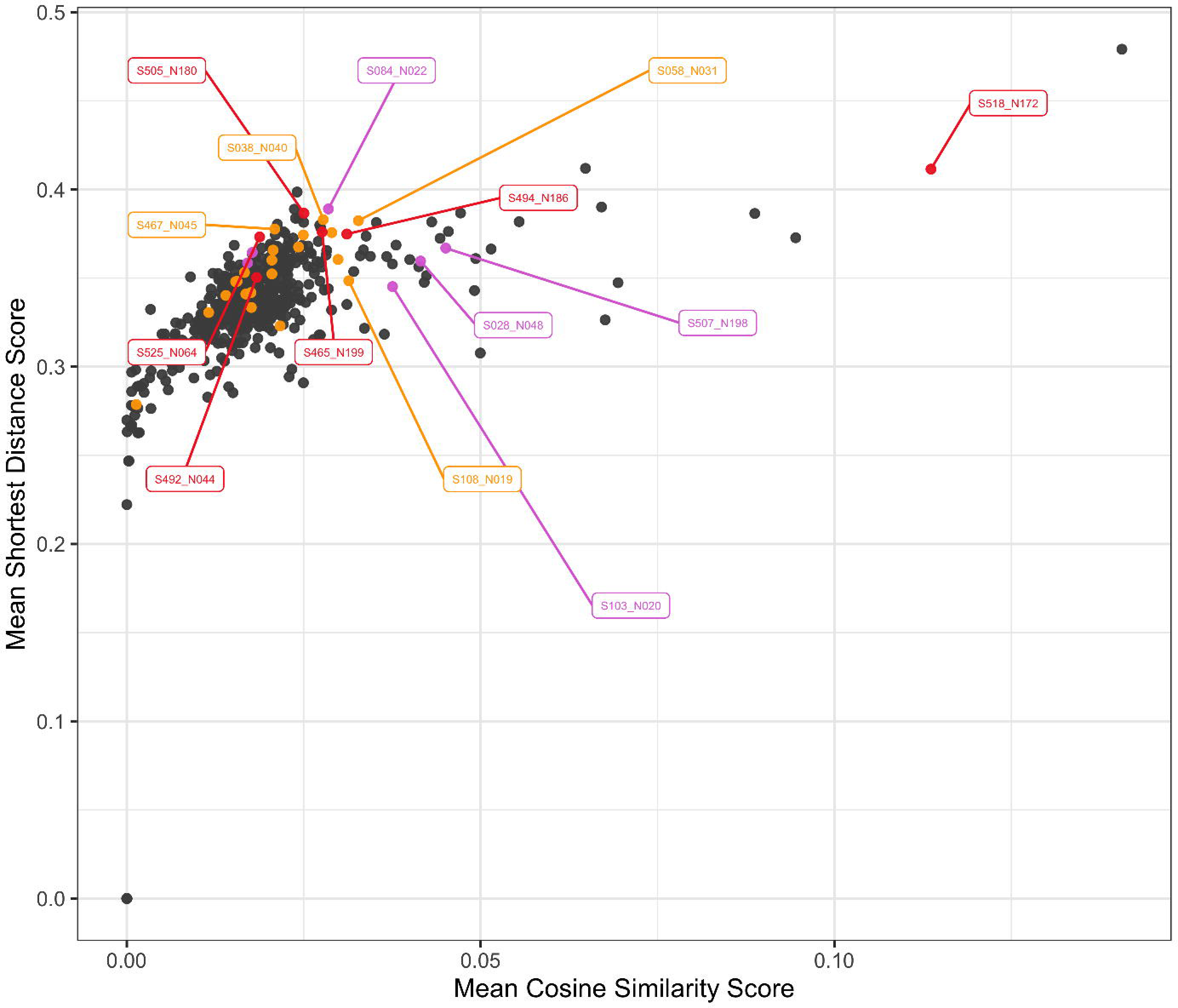
Community proximity to mental health drugs in the network. Multiple communities with enrichment for adiposity associated genes were highly proximal to the mental health drugs. The scatter plot depicts the mean shortest distance scores versus the mean cosine similarity scores for each community and 59 drug vertices (33 antidepressants and 26 antipsychotics). The communities with larger mean shortest distance scores (y-axis) represented groups of genes with more direct paths (shorter paths and stronger edges) between drug vertices and genes in the community. The communities with larger mean cosine similarity scores (x-axis) had evidence for strong network homophily (a high proportion of shared neighbors) with drug-interacting genes (i.e., DrugGenes). Each point represents one of the 536 communities in the network and community labels denote the community identification number followed by the number of gene vertices in that community (e.g., S518_N172 describes community 518 which contained 172 genes). Colors represent FET enrichment p-values (red: below the Bonferroni significance cutoff [p < 9.33x10^-5^], orange: suggestive enrichment [p < 0.01] or pink: communities containing at least 1 monogenic obesity-associated gene).

### Adiposity subnetwork

The interactome captured a diverse and comprehensive set of biological mechanisms, and we were interested in creating a subnetwork of communities in this network that were specifically relevant to the biological processes underlying adiposity and anthropometrics. In Arehart et al., 2025[31] we highlighted two latent factors for abdominal size (F2) and adiposity (F4) which mirror the wADEs affiliated with mental health drugs. From those results, we compiled 5 gene sets (**Tables S4-6**): 1) 730 genes that were significant in GWAS or TWAS of abdominal size (F2 genes); 2) 896 genes that were significant in GWAS or TWAS of adiposity (F4 genes); 3) 17 genes that were characterized in the literature for large monogenic effects on obesity (monogenic genes); 4) 1574 genes representing the union of the first 3 gene sets (F2, F4, or monogenic genes); and 5) 379 refined genes identified by the GRIN random walk with restart algorithm (GRIN-refined F2, F4, or monogenic genes).[46] Fisher’s Exact Tests (FETs) were used to test for enrichment of these gene sets across each of the 536 communities (**Tables S7-8**). We found 6 communities with significant enrichment for at least one of these gene sets (colored red in **Fig. 2**; Bonferroni significant threshold p < 9.33x10^-5^). In addition, 27 communities showed suggestive enrichment (colored orange in **Fig. 2**; p < 0.01), and 10 communities contained a monogenic obesity gene, of particular interest because of their known weight-related genetic effects (colored pink in **Fig. 2**). The union of these communities provided us with an adiposity subnetwork consisting of 33 communities with 2,914 vertices, and 33,372 weighted edges (**Tables S9-10**). Linking the 59 antipsychotic or antidepressant drug vertices into this subnetwork yielded 280 DrugGenes and 1699 drug-protein edges (**Fig. 3**). The 22-gene community(S083_N022) that was most proximal to the mental health drugs (top right point in **Fig. 2**) was not enriched for adiposity-associated genes, and was primarily related to the 13 DrugGenes within the GABA-A receptor, an important ligand-gated ion channel underlying the effects of some antipsychotics and antidepressants.[47–49] Together, these enrichment tests demonstrated that antipsychotics and antidepressants are tightly interconnected with adiposity-associated genes; the weight-related side effects of many mental health drugs may arise because of this network overlap between drug targets and gene products that influence adiposity.

**Fig. 3:**
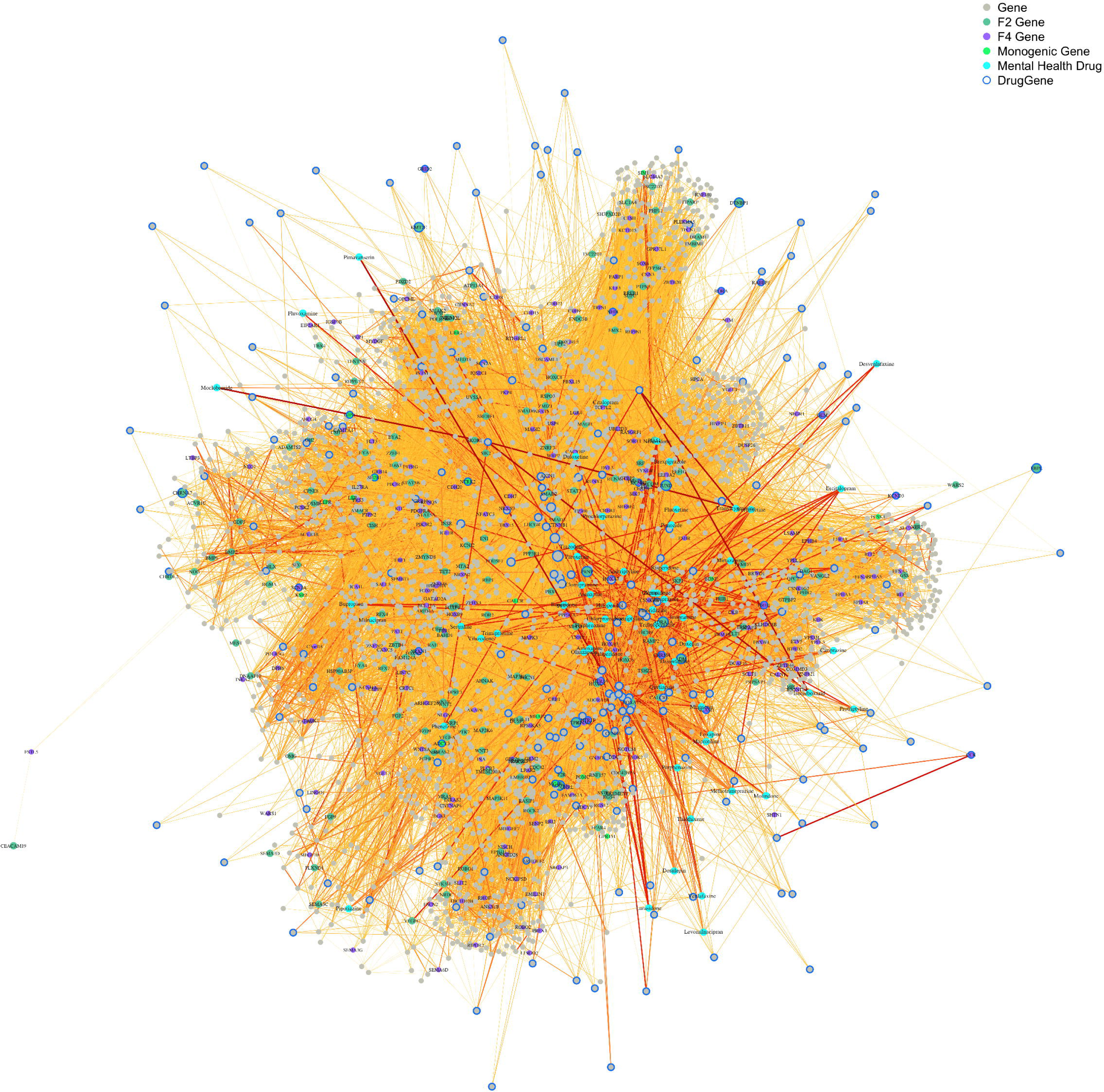
Adiposity subnetwork linked to mental health drugs. Mental health drug vertices were entangled in a highly interconnected subnetwork of adiposity-associated communities. The adiposity subnetwork contained 33 communities with evidence for suggestive enrichment of adiposity-associated genes. This subnetwork consisted of 2914 gene vertices and 33,372 weighted interactome edges. There were 33 antidepressants and 26 antipsychotics connected to 280 drug-interacting genes (i.e., DrugGenes) adding 1699 drug-protein edges.

### DIAMOnD connectivity significance to mental health drugs

We integrated the DIAMOnD algorithm[50] into our analysis as a network pharmacology approach for rank-ordering the interactome’s vertices based on connectivity significance of the adiposity subnetwork (2914 vertices within the 33 communities shown in **Fig. 3**, **Supplementary Data 3**) with the 59 antipsychotic or antidepressant drugs input as seed vertices (**Table S11**). The top 329 iterations identified significantly connected vertices (p < 0.05) based on the DIAMOnD’s statistical test using the cumulative probability of the hypergeometric distribution (**Fig. S8**). Of those 329 DIAMOnD-identified vertices (DIAMOnD-genes), 125 were DrugGenes and 204 did not directly connect to a drug vertex. These top 329 vertices represented a meaningful group of vertices from the 33 adiposity-enriched communities with evidence for mechanistic paths to the mental health drugs. This systematic approach helped us focus on particularly relevant communities in the adiposity subnetwork, with G protein-coupled receptor (GPCR) genes prominent: DIAMOnD-genes were enriched (using IQuery)[51] for GPCR Complete Dataset (116/159 genes; p = 1.38x10^-150^),[52] WP58 - Monoamine GPCRs (33/34 genes; p = 2.12x10^-49^),[53] and INDRA GO:0007188 - adenylate cyclase-modulating G protein-coupled receptor signaling pathway (114/194 genes; p = 4.95x10^-140^).[54] This alternative approach recapitulated what we found with the community gene set enrichment tests described above, which also emphasized the relevance of GPCR signaling as the most pronounced network component connecting mental health drug targets and adiposity genetics. GPCRs encompass 36% of all approved drug targets for a diverse array of indications, including primary MOAs for treating obesity and psychiatric conditions.[55]

### GPCR signaling and the leptin-melanocortin pathway

Multiple analyses triangulated one community in particular, S518_N172 (**Table S12, Fig. S9)**, for its significant enrichment of monogenic obesity genes (p = 2.03x10^-5^), high number of adiposity-associated genes, close proximity to mental health drugs (**Fig. 2**: high mean shortest distance score with drugs [0.41] and high mean cosine similarity score with DrugGenes [0.11]), and prioritized selection by DIAMOnD (**Fig. 4**: 151 of the 172 vertices in this community were included in the DIAMOnD module’s 329 significant iterations). The vertices in this community were largely representative of GPCR signaling processes (via α, β, and γ subunits). These GPCRs identified by DIAMOnD highlighted the central hunger and satiety signaling processes mediated by appetite-regulating neurons in the hypothalamus:[56–59] POMC (D128 [selected in DIAMOnD’s 128^th^ iteration]) neurons activate MC4R (D104) after PCSK1/PCSK2 (D632/D301) cleaves α-MSH to suppress appetite, while AGRP (D201) neurons function to stimulate appetite. MRAP2 (D279) modulates MC4R (D104) and ADCY3/ADCY5/ADCY9 (D113/D69/D174) facilitate MC4R Gs signaling in which the activation of adenylyl cyclase leads to increased cAMP, relieved inhibition of PKA, and the phosphorylation of downstream targets. In addition, multiple RGS proteins RGS4/RGS12 (D150/D282) and ARRB1/ARRB2 (D115/D113) are notable for their roles in regulating GPCR activity.[60,61] TPH1 (D41) and DDC (D35) are important to the biosynthesis of serotonin and dopamine, neurotransmitters that shape the activity of these brain circuits and, in the case of peripheral serotonin, contribute to increased lipid storage.[62] In this community, peripheral hormone signaling from the gut via GIPR/GLP1R (D242/D243) and the stomach via GHRL (D170) were also implicated, along with fatty acid receptor FFAR4 (D67) which are all involved in appetite regulation, metabolic homeostasis, and insulin signaling. In summary, many proteins that are central to hypothalamic signaling and the leptin-melanocortin pathway were identified within this community.

**Fig. 4:**
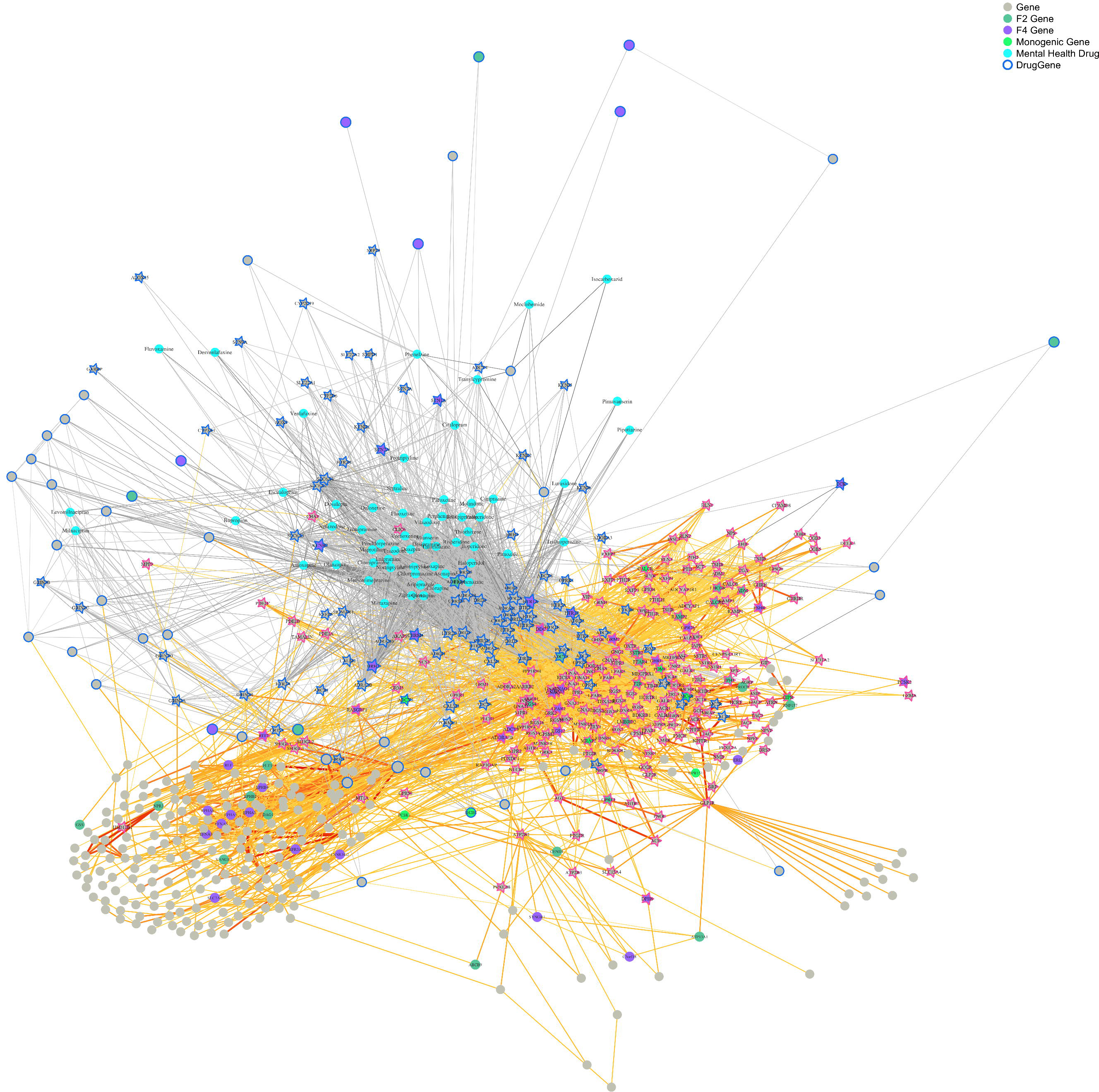
Connectivity significance to mental health drugs. The top 329 significant iterations of DIAMOnD-identified vertices primarily belonging to 6 communities in the adiposity subnetwork: S518_N172 (151/172 vertices), S028_N048 (48/48 vertices), S507_N198 (10/198 vertices), S103_N020 (7/20 vertices), S108_N019 (6/19 vertices). These 6 communities’ interactions are shown along with their links to the 59 drug vertices (the seed vertices for the DIAMOnD algorithm). The top 329 iterations identified significantly connected vertices (p < 0.05) which are represented as star-shaped vertices outlined in pink (or blue for the DrugGenes).

The DIAMOnD-prioritized genes in **Fig. 4** implicated multiple monoamine and peptide GPCRs that amplify the same signaling cascades as the hunger and satiety pathways[63] as a potentially central mechanism by which mental health drugs cause wADEs. While GPCRs are fine-tuned to use different couplings of α, β, and γ subunits to spur a variety of downstream cell behaviors[52], our interactome illustrated the overlapping use of α subunits and β/γ subunits among GPCRs in this community (DIAMOnD identified 13 α subunits and 1 γ subunit within the first 97 iterations). DIAMOnD-identified GNAS (D66) and GNAL (D62) were primary α subunits of interest given their role as messengers for the G_s_ classification of GPCRs that are core to the satiety response such as MC4R, GLP1R, and GIPR.[64] The G_s_ signal transduction activates adenylyl cyclase which converts ATP to cAMP and results in the activation of PKA. In contrast, GPCRs operating through G_i/o_ complexes have opposing biological effects through the inhibition of adenylyl cyclase. Our results unveiled the hypothesis that mental health drugs alter the levels of cAMP (through acting as agonists/antagonists for G_s_ GPCRs and/or G_i/o_ GPCRs), converging on the same intracellular signaling mechanism as MC4R in hypothalamic neurons. The overlapping use of α and β/γ subunits among GPCRs that bind these drugs illustrates how the same proteins often participate in more than one pathway, and various extracellular signaling events (e.g., hormones or drugs interacting with G_s_ or G_i/o_ GPCRs) could result in similar intracellular responses as these signals are integrated.

For example, the 5-hydroxytryptamine receptor 7 (*HTR7*), a prominent DrugGene in our network targeted by 38 of the 59 antipsychotic or antidepressant drugs, is known to have primary coupling with the same G_s_ complexes as MC4R (GNAS and GNAL; **Fig. 5a**),[52] and is highly expressed in the hypothalamus.[65,66] While the direction of effect is unspecified for many of the interactions between mental health drugs and HTR7, DrugCentral[67] highlighted 15 of these interactions as antagonist actions (e.g., antidepressants including amitriptyline, vortioxetine, and antipsychotics including olanzapine, clozapine, lurasidone, brexpiprazole, and haloperidol). These antagonizing interactions in hypothalamic neurons could plausibly counteract the G_s_ signaling that is core to the MC4R satiety response, leading to increased eating and drug-induced weight gain.[68,69] In a similar manner, GPCRs that primarily signal through G_i/o_ α subunits could also contribute to this integrated signaling hypothesis. For example, the dopamine D2 receptor (DRD2) interacted with 47 of the 59 antipsychotic or antidepressant drugs and primarily couples the G_i/o_ family of α subunits (GNAI3, GNAI1, GNAO1, GNAZ; **Fig. 5b**).[52] A number of these drug-protein interactions described in DrugCentral[67] and the DGIdb[70] are classified as agonist actions (including brexpiprazole, aripiprazole, vilazodone, cariprazine). *DRD2* is moderately-to-highly expressed in the hypothalamus,[71,72] and agonists of DRD2 could lead to decreased cAMP levels, reduced PKA activity, and dampened satiety signaling in hypothalamic neurons. Given the number of GPCRs and mental health drugs interwoven throughout the network (**Figs. 3-4**), it is challenging to trace the possible outcomes of signaling trajectories from agonist versus antagonist drug-protein interactions. Different drugs may activate or interfere with the same gene product with differing magnitudes in opposing cell types (orexigenic and anorexigenic neurons) which complicates direct interpretation, but these results generate a broader hypothesis for the importance of understanding how GPCR signals are integrated in the hypothalamus.

**Fig. 5:**
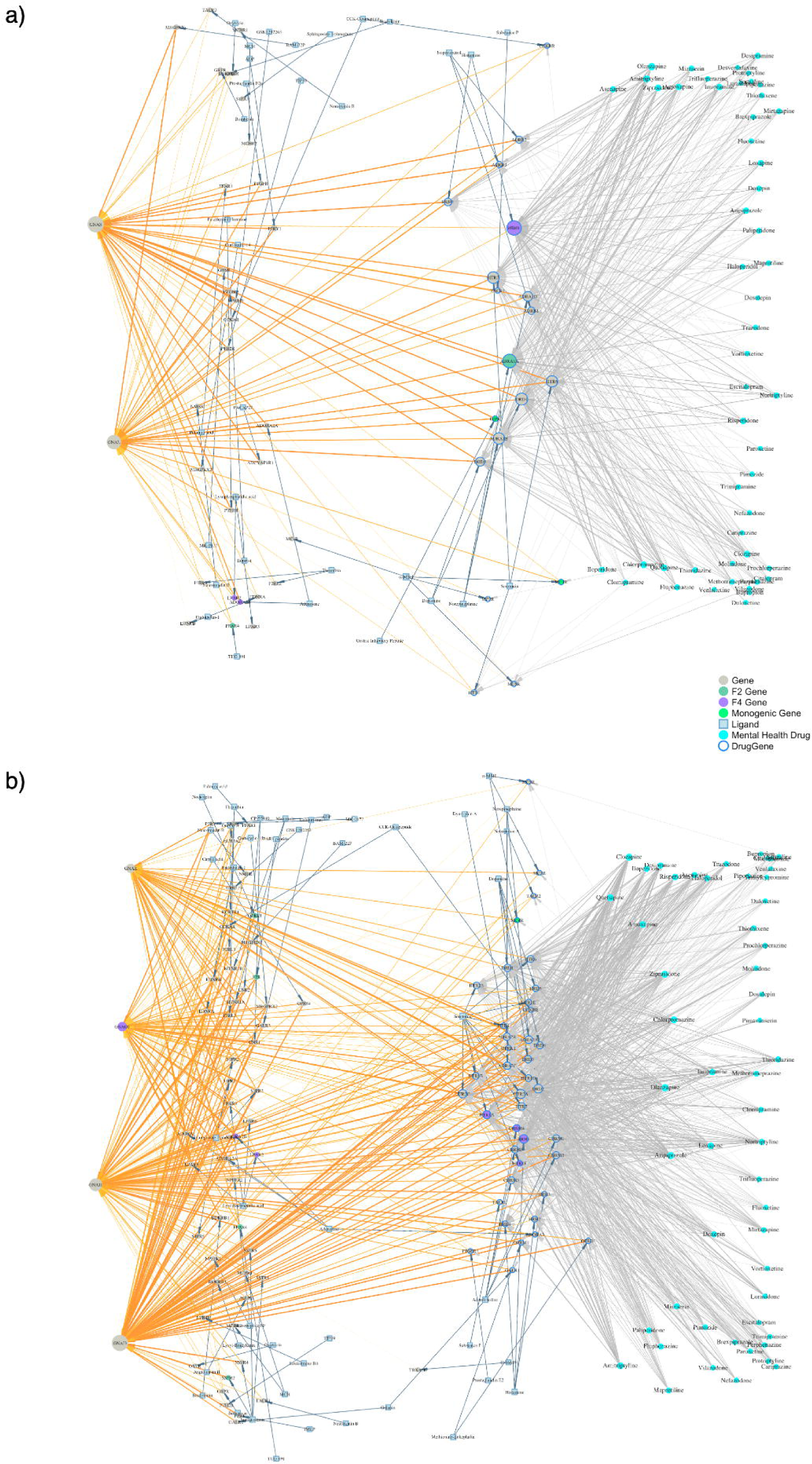
Overlapping GPCR signaling between hunger/satiety mechanisms and DrugGenes. GPCRs were emphasized for their proximity to the mental health drugs and many of the DrugGenes in the network primarily signaled through the same G_s_/G_i/o_ pathways as MC4R. Panel a illustrates GPCRs that couple with G_s_ complexes, and panel b illustrates GPCR that couple with G_i/o_ complexes. Edges between vertices were defined using the Inoue et al., 2019 GPCR Complete Dataset, including interactions with high-confidence relative intrinsic activity (LogRAi ≥ −1).[52] Drug-Protein interactions are shown in grey, ligand-GPCR interactions are shown in blue, and GPCR-G_α_ interactions are shown in orange with darker coloring corresponding stronger coupling (larger LogRAi).

While GPCR α subunits target key effector proteins, the β and γ subunits also have important signaling roles inside the cell. The β and γ subunits form a dimer and target effector proteins of their own once GPCRs are activated, such as sodium (SCN2A/SCN1A/SCN3A identified by DIAMOnD: D58/D59/D60) and calcium ion channels (CACNA1H/CACNA1G/CACNA1I; D370/D371/D372).[73,74] Opening K⁺ channels (e.g., KCNJ6/KCNJ5; D46/D124) could lead to hyperpolarization of neurons and inhibit neuron firing, and inhibiting Ca²⁺ influx could reduce neurotransmitter release.[75] Both of these mechanisms could lead to dampened signaling of hunger- or satiety-promoting neurons contributing to wADEs. Our results emphasize the importance of GPCRs as crucial cellular components underlying both drug targets and the etiology of adiposity. Further untangling this signaling overlap would be a promising area of research for improving the development and target specificity of mental health drugs.

### Random walks between adiposity genes and mental health drugs

We applied a random walk with restart (RWR) framework to determine which edges and vertices in the network bridged the adiposity-associated genes to the mental health drugs, with the goal of identifying key biological pathways. These random walks provided an effective strategy for controlled exploration of network connectivity to highlight the central or most commonly traversable network paths between adiposity-associated genes and mental health drugs (**Fig. S10a**; see methods). The resulting RWR network (1609 vertices and 4942 edges) summarized the paths that were visited with edge weights and node degrees representing traversal frequency (**Fig. S11-12**). To improve pathway interpretability, we integrated information from OmniPath[36] describing signaling directionality (source and target vertices) and action (stimulatory, inhibitory, neither, or both) throughout the RWR network.[36] In addition, we applied an intuitive RWR traffic score (**Fig. S10b**; see methods) to prioritize edges with high participation across the random walks – edges between nodes with high weighted degrees (transit hubs) and edges with high traversal frequency (bottlenecks). Such highly trafficked nodes and edges (i.e., with high betweenness) represent hypotheses of mechanisms underlying why antidepressants and antipsychotics induce weight changes. PRKACA, PRKCA, FYN, EGFR, ADRB2, SRC, and CALM1 had the highest sums of traffic scores in the RWR network (**Table S13**). The convergence of these random walks on kinases PRKACA and PRKCA recapitulated the importance of the G protein activation pathways discussed above. We additionally highlighted a frequently traversed topological route in the RWRs involving calcium signaling, calmodulin binding (CALM1/CALM2/CALM3; also identified in significant DIAMOnD iterations D187/D264/D259), and calcitonin/amylin binding (CALCR; D238). All 48 of the vertices in community S028_N048 (which contained F2-associated genes: ADM, CALCRL, RAMP2, LHCGR, F4-associated gene: FSHR, and monogenic-associated gene: CALCR) were also independently identified by DIAMOnD (**Fig. S9**). Patterns of Ca^2+^ within adipocytes and hypothalamic neurons regulate energy metabolism through the P38-MAPK, Calmodulin, IP3- Ca^2+^, cAMP-PKA, and Wnt-Ca^2+^ signaling pathways.[75] For the key edges with high traffic scores, we used enrichment tests in IQuery[51] of their 2-step ego networks to relate their proximal RWR network surroundings to biological functions. This identified numerous potential mechanisms by which the 59 mental health drugs may lead to wADEs. Below, we outline 3 vignettes to explore a few of these bottlenecks in more detail.

### PI3K-Akt Signaling

The edge between EGFR and MET had a traffic score of 0.102 and its 2-step ego network (**Table S14**) included 295 protein vertices which were enriched in IQuery[51] for WP4172 - PI3K-Akt Signaling (51/339 genes; p = 2.68x10^-24^)[53] and SIGNOR - PI3K/AKT Signaling (35/144 genes; p = 1.36x10^-23^).[76] 244 of those 295 vertices were adiposity-associated genes (including monogenic obesity genes) illustrating the overlap between PI3K/AKT signaling and adiposity-associated gene sets (**Fig. 6**). The PI3K-Akt signaling pathway is vital to leptin signaling, insulin signaling, and thermogenesis.[77] Generally, this pathway is a key regulator of cell growth, proliferation, and survival and has important functions in skeletal muscle, adipose, liver, hypothalamus, pancreas tissues.[78] Activation of growth factors, RTKs, cytokines, and GPCRs leads to the conversion of PIP2 to PIP3 and the recruitment of AKT. AKT proteins have many downstream effectors that are involved in metabolism and energy homeostasis. Our RWR analysis emphasized many genes in the PI3K-Akt pathway.

**Fig. 6:**
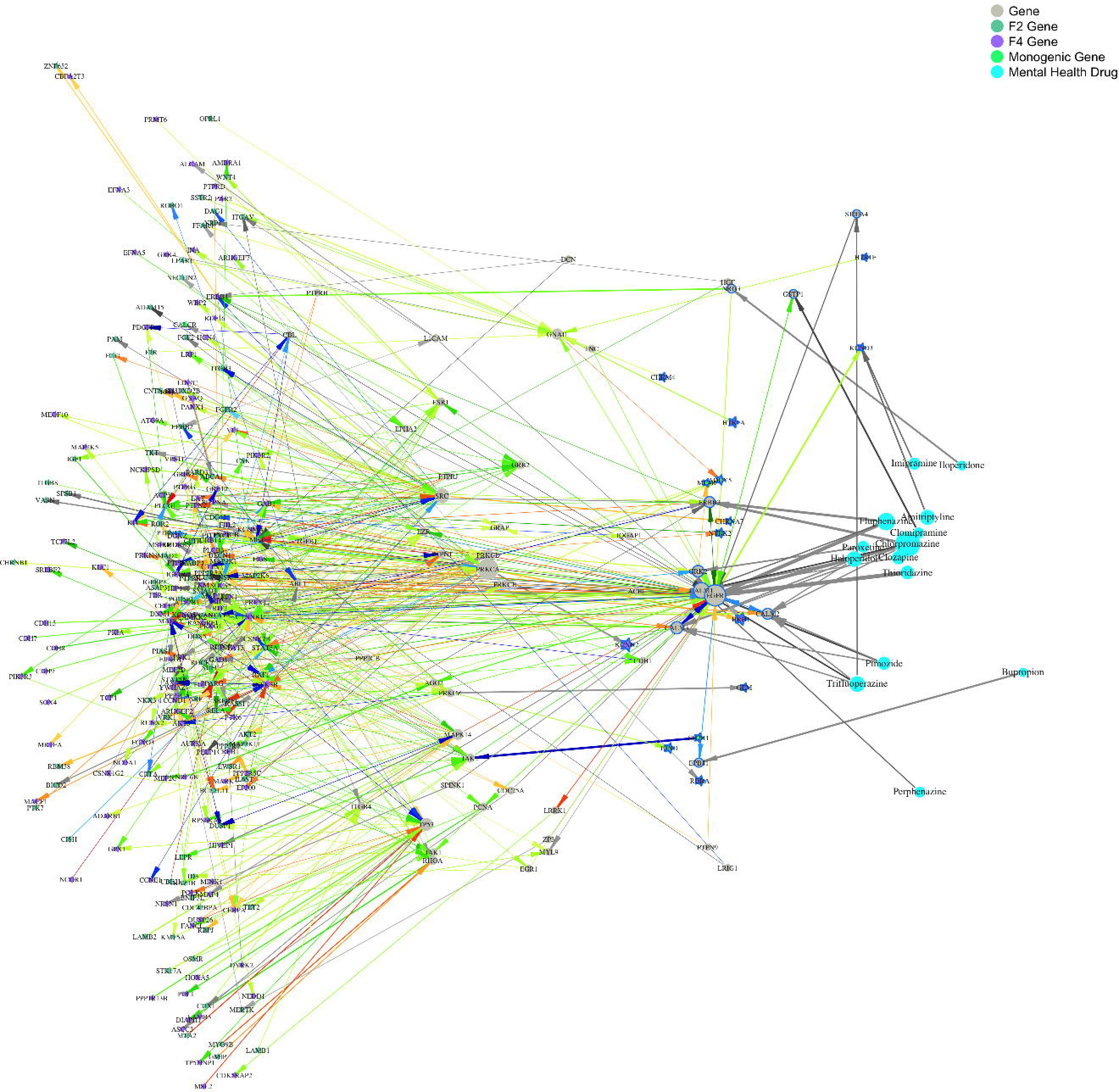
Random walks with restart highlighted the PI3K-Akt signaling pathway. From the full RWR network (Fig. S11), the edge between EGFR-MET unveiled a 2-step ego network that was significantly enriched for the PI3K-Akt pathway. Arrows and colors indicate the OmniPath directionality (or bidirectionality) from source to target vertices and type of action (green for stimulatory, red for inhibitory, grey for neither, and blue for both). Upstream components included growth factors, RTKs, GPCRs, and cytokine receptors along with AKT2, AKT3, and downstream proteins signaling for metabolism, cell cycle progression, and cell survival.

### Leptin signaling

Our RWR analysis also identified genes central to the leptin signaling pathway (**Fig. S18**), such as the JAK2-SH2B1 edge (traffic score of 0.189). The 2-step ego network for this edge (**Table S15**) included 75 protein vertices, 66 of which were among the starting vertices (associated with F2, F4, or monogenic obesity). These genes were enriched in IQuery[51] for WP2034 - Leptin signaling (13/71 genes; p = 2.57x10^-14^)[53] and SIGNOR - Leptin Signaling (11/41 genes; p = 4.21x10^-14^).[76] The directionality of the arrows derived from OmniPath illustrates the flow of interactions that could sequentially propagate from the mental health drugs toward the JAK2-SH2B1 edge and then connect to many adiposity genes. Leptin is a signaling hormone produced by adipose tissue which binds neurons in the hypothalamus to induce appetite-suppressing effects. When leptin binds its receptor, it activates JAK2 leading to the recruitment of STAT3 which translocates into the nucleus to increase *POMC* expression and decrease Npy/Agrp expression (leading to anorexigenic effects).[79] *SH2B1* is a monogenic obesity associated gene and an important molecule for regulating the JAK2-STAT3 pathway and leptin sensitivity.[80]

### Adipogenesis

High traffic edges between drugs and adiposity genes also unveiled potential mechanisms by which mental health drugs’ molecular interactions lead to changes in the accumulation of adipose tissue (**Fig. S19**). The RWR network edge between NEDD4L-SMAD3 highlighted the many interactions surrounding PPARG and CEBPA relating to adipogenesis. The 2-step ego network for this edge (**Table S16**) included 221 protein vertices and 185 of them were associated with F2, F4, or monogenic obesity. IQuery[51] results highlighted enrichment for WP236 - Adipogenesis (20/131 genes; p = 6.13x10^-12^)[53] and SIGNOR – Adipogenesis (15/27 genes; p = 9.42x10^-18^).[76] The expression of *PPARG* and *CEBPA* is central to preadipocyte commitment and differentiation; surrounding these core proteins, an orchestration of multiple signals and transcription factors determines the transition from precursor stem cells into mature adipocytes.[81] Within this ego network, we identified transcription factors and pathway modulators in the WP236 - Adipogenesis pathway including CREB1, MEF2C, MEF2D, SREBF1, NCOA1, RORA, NCOR1, TGFB1, IGF1, NR3C1, SMAD3, CTNNB1, STAT3, RBL2, and PCK1. Many of the drugs linked into KCNH2 which is an adiposity-associated DrugGene with a central role in the potassium channel regulation of incretin secretion (e.g., GIP and GLP-1).[82] KCNH2 had the 10^th^ highest sum of RWR traffic scores, and its intersecting roles with incretin regulation and mental health drug interactions demonstrated both its relevance and its draggability.

## Discussion

We aimed to deconvolve the mechanistic links between mental health drugs and their adverse weight-related side effects. From our biological interactome, we identified links between adiposity-associated genes and wADE-inducing antidepressant and antipsychotic drugs. We implemented multiple network modeling approaches (network clustering and proximity measures, DIAMOnD connectivity significance, and RWRs), which converged on plausible hypotheses involving GPCR signaling, calcium signaling, PI3K-Akt signaling, leptin signaling, and adipogenesis. We identified links to both the monogenic forms of obesity, such as genes that are core to the leptin melanocortin pathway, as well as polygenic forms of obesity, with genes that operate across more complex signaling mechanisms. A central finding of our analyses, however, is that of overall crosstalk between biological pathways – mental health drugs may have weight-related adverse side effects because the proteins they target are also integrated in pathways implicated in adiposity. wADEs are a central hindrance to psychiatric treatments and disentangling the mechanisms of how they influence weight changes could help improve treatments and design new therapies.

Our study inherits the potential sources of error underlying the datasets that were incorporated along with the limitations underlying the analytical tools which we applied. Our interactome, like others,[33,34,36–38,40] was not cell-type or tissue specific which limited our ability to interpret the sequences of biological interactions for the RWR analysis. In addition, for the RWR analysis we were limited to a smaller subset of edges in OmniPath with identified directionality and inhibiting/activating effects. Improving the full interactome to model trait-specific expression patterns, edge directionality, and type of action (e.g., a directed and weighted multiplex) would greatly improve the interpretability of our outcomes. Nonetheless, this exploratory analysis generated detailed functional hypotheses for future validation that may facilitate drug design and enhance drug-target discovery efforts.

Beyond our specific application regarding adiposity and mental health drugs, this analysis presents a broadly applicable methodological framework for exploring the molecular underpinnings of complex diseases. We have shown that it is possible to identify communities of interacting genes that are proximal to a class of drugs, identify the pathways that involve genes in those communities, and annotate interconnected communities that are enriched for trait-associated genes. Because these drugs have undergone rigorous approval processes with randomized control trials to establish causal effects (considering both targeted diseases and off-target side effects), linking drug targets and associated genes can improve confidence to triangulate key biological mechanisms that are druggable and have causal implications within a broad landscape of genomic associations. This generates new opportunities to apply our framework (using the same underlying interactome and methodological pipeline) to other observed pairings in which an adverse drug event mirrors a trait of interest.

## Methods

### Experimental Design

We devised a robust analytical framework to interrogate potential mechanistic paths between drugs and their adverse side effects. Here, we focused on mental health drugs and their adverse side effects relating to weight gain and weight loss, using a network modeling approach to identify putative mechanisms. We began by constructing a biological interactome encompassing a robust collection of molecular interactions. We then clustered this network, and after performing a series of enrichment tests for adiposity-associated genes we identified a subset of communities with functions that appeared relevant to adiposity. Multiple databases were used to link antipsychotic and antidepressant medications into our interactome, and we identified communities with high proximity to the mental health drugs. We then evaluated connectivity significance to the mental health drugs and rank ordered the vertices in the adiposity subnetwork. Finally, we performed random walks between adiposity-associated genes and the mental health drugs to prioritize network edges with high betweenness. These analyses enabled us to zoom in on the components of the interactome that were most relevant to our research question and emphasized specific pathways with evidence for linking mental health drugs to their wADEs. In the sections below we provide detail for how we constructed an improved biological interactome and performed multiple analyses to explore the space between mental health drugs and the biology of adiposity.

### Improved Biological Network (Interactome)

The core network used in our analysis was constructed using 7 databases that were each curated from a variety of sources with differing criteria (**Table S1**). We used these databases’ protein-protein interactions (PPIs) or gene-gene interactions (GGIs) to build a meaningful network representation of human biological processes consisting of proteins/genes (vertices) and their interactions (edges). Through combining these databases, we were able to leverage information from multiple biological layers to capture physical PPIs, biological signaling, and genetic co-expression modules in the brain. The 5 primary databases included the STRING[33] (v12.0) physical subnetwork with scored links between proteins, the InWeb_InBioMap (aka InWeb_IM)[34] scored human protein–protein interaction network, the BioGRID[35,37] MV-Physical (v4.4.236) physical multi-validated human protein-protein interaction network, the OmniPath[36] literature-curated human signaling interaction network, and HIPPIE[40] v2.3. The 2 auxiliary databases included the Gandal et al., 2018 neuronal co-expression modules (GNCEM)[39] with module-membership strengths (kME values) and the Harmonizome[38] dataset which aggregates biological relationships across a broad range sources.

Within the comprehensive STRING database, we selected the subnetwork of direct (physical) associations, thereby excluding the functional associations (participation in the same pathway) and filtered out edges with low confidence scores (< 0.15). The InWeb_InBioMap scores ranged from 0 to 1, and the kneedle algorithm[83] was implemented (via the kneedle R package; sensitivity = 1000) to identify the distinct knee-point in the ranked interaction scores. The algorithm identified 0.112 as the knee-point, and this threshold was used to identify the subset of edges – 88,486 of the 450,083 InWeb_InBioMap edges – which had a confidence score greater than 0.112 (see criteria below). The GNCEM primary co-expression modules (CD1, CD5, CD10, CD13, CD4, CD11, CD12, CD2)[39] were incorporated and each edge weight was defined as the pairwise sum of kME module membership scores. These scores were rescaled to range from 0 to 1. BioGRID, OmniPath, and Harmonizome edges did not include confidence scores and were all assigned an intermediate weight of 0.5.

The first criterion in constructing the network was that an edge had to be identified by at least 1 of the 5 primary databases (STRING, BioGRID, InWeb_InBioMap, OmniPath, or HIPPIE), unless it was identified by both auxiliary databases (Harmonizome and GNCEM). We also removed edges that were only identified in STRING with low-to-moderate confidence scores (using STRING-suggested medium confidence cutoff < 0.7).[33] Similarly, we removed edges that were only identified in InWeb_InBioMap with low scores (below the knee-point of 0.112). This threshold was similar to the 0.154 cutoff used in other studies, including the InWeb_InBioMap database.[84] Following the HIPPIE documentation, edges with solitary identification in HIPPIE and scores less than 0.73 (high confidence - third quartile threshold) were removed. Finally, any edges with scores below these respective thresholds that were only identified by 2 of the scored databases (STRING, InWeb_InBioMap, and HIPPIE) were also removed. In the end, 408,883 of the 5,314,131 considered edges were included in our biological network (**Table S1**) and the edge weights were summed across the databases. Post summation, these scores were rescaled to be between 0 and 1. This final network contained 17,293 vertices representing gene products (e.g., proteins), and the strength of redundancy across databases was reflected in the final edge weights (81% of the edges were identified by at least 2 of the 7 databases) (**Fig. S2**; **Supplementary Data 1**). Our interactome thus aggregated evidence from high-throughput protein interaction assays, curated pathways, text mining of published scientific literature, co-expression modules, and interlogs.

### Network Clustering

In order to identify clusters of tightly interconnected genes within our network, we compared multiple community-detection techniques. We evaluated the MONET-K1, MONET-M1, MONET-R1,[43] Big Clam,[85] leading eigenvector,[86] Louvain,[87] and Leiden[88] algorithms.[89] Each of the non-MONET-based methods failed to return modules of reasonable sizes (i.e., they returned very large modules with thousands of genes), and these large modules were less useful for our goal of gaining biological insights from specific parts of the network. The DREAM challenge, however, required each method to produce modules with less than 100 genes which was more suitable for our intended analyses.[44] In order to compare the MONET K1, M1, and R1 methods we ran each using the standard parameters described in the documentation found at https://github.com/BergmannLab/MONET. We visualized each method’s defined modules containing randomly selected sets of 5 genes to compare the interconnectedness within and between the modules (**Fig. S4**). The K1 method often returned modules with vertices that had no connections to other vertices in the module, and the R1 method generally produced modules that failed to appear topologically distinct. The M1 method, however, illustrated groups of vertices that tended to show strong patterns of interconnectedness – even when visualized in a broader context of surrounding network vertices. Thus, we decided to use the MONET-M1 algorithm, which provides fine-tuning of vertex cluster assignments based on an ensemble consensus across multiple modularity-optimized algorithms/runs. In the DREAM challenge,[44] the M1 algorithm performed well on biological networks similar to ours (STRING, InWeb_InBioMap, and OmniPath) and was easy to implement using the command-line tool MONET.[43] We identified 1029 modules ranging in size from 2 to 98, and these modules contained 15,390 of the 17,293 network vertices. The maximum module size threshold of 100 was an original constraint in the DREAM challenge and was a fundamental parameter to the design and evaluation of the M1 clustering method. The median and mean module sizes were 10 and 14.96, respectively (**Fig. S5**).

While this fragmentation of the network yielded a highly granular set of modules, many of those modules were too small for downstream analyses (e.g., enrichment tests). Simulations for Fisher’s exact tests suggested that in order to characterize network modules for their enrichment of relevant gene sets (curated pathways or ontologies, or associated genes from GWAS or TWAS), it would be optimal to have modules with at least 10-30 vertices (**Fig. S6**). Thus, we designed a greedy approach to consider merging smaller modules with high measures of network similarity and proximity (i.e., merge two modules with many common neighbors and relatively short paths among their sets of vertices). We calculated the cosine similarity matrix (17,293 x 17,293) for all vertices using the network’s weighted adjacency matrix (denoted A hereafter). Entries in the resulting symmetric cosine similarity matrix ranged from 0 to 1 representing the similarity of pairs of weighted vertex-feature vectors. Intuitively, a large cosine similarity value (closer to 1) indicated that two vertices had many shared neighbors while a small cosine similarity value (closer to 0) implied they had few in common (while accounting for overall vertex degrees). We also computed a symmetric matrix of shortest distance scores (17,293 x 17,293) using Dijkstra’s algorithm.[90] The original edge weights in A represented the confidence or strength of an interaction between 2 vertices, so before calculating the shortest distances between each pair of vertices we reversed the weights (1 – A). We then applied Dijkstra’s algorithm via the igraph[89] distances function in R and converted the resulting weighted path lengths into scores ranging from 0 to 1, such that values closer to 1 signified closer network proximity in terms of path tracing: 1 / (1 + ‘sum of shortest path weights’). The cosine similarity and shortest distance matrices were then used to calculate the mean similarity and mean shortest distance matrices for all modules identified by MONET-M1 (two, 1029 x 1029 symmetric matrices; **Fig. S13**). For example, the (2,5)^th^ entry of one of these symmetric matrices represented the mean proximity/similarity score of vertices in the 2^nd^ module with vertices in the 5^th^ module. A pair of modules with high mean cosine similarity and shortest distance scores reflected evidence of network homophily and proximity, thereby indicating their candidacy for being merged into a joined community as estimated using the root mean square (RMS), also known as the quadratic mean (or the generalized mean with p = 2). The RMS provided a more meaningful mean of pairwise cosine similarities between two modules through emphasizing the influence of larger values. In this application, the RMS was generally similar to the arithmetic mean, however, it increased the consideration of particularly similar vertices from pairs of modules. We only considered merging a pair of modules if they exhibited high scores for mean similarity (**Fig. S14**) and mean shortest distance (**Fig. S15**), and 6,743 out of 528,906 pairs of modules (1.27%) met the criteria of having mean cosine similarity ≥ 0.0527 and mean distance score ≥ 0.3754 (thresholds identified using the kneedle algorithm).[83] The 6,743 pairs of modules with high similarity scores and distance scores (**Fig. S16**) were rank-ordered by a combined Euclidian score, sqrt(similarity^2^ + distance^2^), and assessed for potential merging in 3 phases based on community size. First, all 6,743 pairs of similar/proximal modules were considered for a merge if either of the modules was sized < 10 based on their ranking of combined Euclidian scores. In the next phase, we considered merging modules if either was sized < 20, and finally, we considered merging modules if either was sized < 30. In each of the 3 stages all 6,743 pairs of modules were considered in rank order, and merges were prevented if the sum of both module sizes was ≥ 200. In the instance of a successful merge, the two original modules were replaced by a combined community encompassing the union of their vertices. This ultimately resulted in 536 communities (median size = 11.50, mean size = 28.71, range = 2-199) for our downstream analyses. The size of these 536 communities remained small enough to preserve the desirable fine-grained structure of network partitioning (interquartile range of community size: 50-88 [**Fig. S5**]), and they were more adequately sized for performing enrichment tests compared to the original 1029 modules.

### Adiposity Enrichment Tests

We were interested in empirically isolating a sub-network of communities within the network that were pertinent to weight-associated genes. In order to test for enrichment of adiposity-associated genes within each community we first had to define sets of weight-associated genes. In previous work,[31] we constructed a 4-facter genomic structural equation model to disentangle the genetics of adiposity and anthropometrics across the lifespan. This model thoroughly characterized the genomic and transcriptomic associations for birth size (F1), abdominal size (F2), body size/composition (F3), and adiposity (F4). We discovered that F2 carried predictive utility for endocrine, metabolic, and circulatory disorders (e.g., type 2 diabetes, hyperlipidemia, and hypertension) and had significant positive genetic correlations with the components of MetSyn, substance use, internalizing behaviors, and frailty. In addition, we found that F4 was the most central genetic factor predisposing individuals to adverse health outcomes with many phenotypic associations, strong genetic correlations with affiliated traits, distinct gene enrichment for neuronal tissues, and overlap with psychiatric drugs targets. The gene set for F2 relating to abdominal size contained 730 genes and the gene set for F4 relating to adiposity contained 896 genes that were significant in their respective GWASs or TWASs and present in the interactome. To complement the polygenic signal captured by F2 and F4 for abdominal and common obesity, we compiled a list of 17 monogenic obesity genes from the literature that have been well characterized for their large effects often resulting in rare, early-onset, and severe forms of obesity (**Table S5**).[58,91] The union of these 3 gene sets resulted in 1574 genes (F2, F4, or monogenic genes).

We were interested in using a network-based approach to refine this large gene set and emphasize genes that were more likely to be true positives (the associated SNPs from our GWASs could have prioritized incorrect genes in close proximity, or the fine-mapping of credible sets from our TWASs could have incorporated false positives due to the underlying correlation structure of gene expression weights and linkage disequilibrium). Thus, we applied GRIN[46] which explores a biological network’s topological structure to compare the interconnectedness of candidate genes to random genes in order to filter out those that are more likely to be false positives. GRIN’s underlying random walk with restart approach ranks the provided genes to produce a subset of “retained GRIN Genes” that (based on network evidence) are more likely to truly contribute to the trait of interest. We applied GRIN using our interactome containing 17,293 vertices and 408,883 undirected weighted edges (constructed a single layer multiplex using RWR_make_multiplex with two edges for each source/target pairing). We input the list of 1574 seed F2, F4, or monogenic genes along with the default GRIN parameters for -- restart (value = 0.7) and --tau (value = 1). This resulted in a smaller set of 379 refined genes identified by the GRIN random walk with restart algorithm (**Table S6**). Ultimately, we aggregated 5 gene sets for our network analysis: F2, F4, monogenic, F2/F4/monogenic, and GRIN-refined-F2/F4/monogenic (**Tables S4-6**).

Fisher’s Exact Tests were used to test for enrichment of these 5 gene sets across the 536 communities in the interactome. We assessed significant enrichment using the Bonferroni-corrected significance threshold (p < 9.33x10^-5^) and suggestive enrichment using a more lenient threshold (p < 0.01). We also identified communities with at least 1 monogenic obesity gene, since the large-effect and penetrant nature of those genetic associations could conceivably warrant further investigation in the context of our research question. The union of communities that were significant, suggestive, or contained a monogenic gene provided us with an adiposity subnetwork consisting of 33 communities with 2,914 vertices, and 33,372 weighted edges. These key communities of the sub-network were then visualized using the igraph R package[89] with spatial clustering of vertices belonging to the same community. Next, we used Network Data Exchange Integrated Query (NDEx IQuery)[51] to characterize the biological function and pathway-based interpretation of these communities. This tool provided an annotation framework via gene set enrichment tests to identify the most plausible biological processes affiliated with each community. We tested each community against the IQuery curated pathways and INDRA gene ontologies[54] to uncover etiologies that are fundamental to the biology of weight gain or weight loss.

### Linking Drugs to the Biological Network

We began by identifying drugs relevant to our research question through compiling a comprehensive list of mental health drugs. Indications for drugs were identified using the ensemble MEDication Indication resource (MEDI-C)[45] containing 38,378 high precision drug-indication pairs. The PheWAS R package[92] enabled mapping of the indications (ICD10CM codes) to phecodes with corresponding phenotype domains, and we subset to drugs within the phecode group “mental disorders.” We used Drugbank[93] IDs to unify drug identifications when merging across databases, and the DrugBank’s listing of Anatomical Therapeutic Chemical (ATC) classifications to refine the list of drugs extracted using MEDI-C. There were 5 levels of ATC classifications, and we used Level 4 which goes beyond pharmacological subgroup and further categorizes drugs based on chemical structure and specific pharmacological properties. We accumulated the interactions for 59 mental health drugs with ATC classification of “antidepressants” or “antipsychotics” (33 antidepressants, and 26 antipsychotics). These were the two largest ATC classifications and the most relevant to our question regarding mental health drugs with common side effects relating to changes in weight. The ON-label SIDE effectS resource (OnSIDES, data release 2.1.0-20240925)[94] was used to identify wADEs for the drugs in the network following the same protocol outlined in Arehart et al., 2025.[31] Of the 59 antidepressant or antipsychotic medications, 43 had a wADEs (e.g., increased/decreased appetite, or weight increased/decreased) 7 did not have a wADE listed, 9 were missing from OnSIDES (unknown wADE). These drugs and annotations are described in **Table S3**.

Drugs are known to interact with both on-target and off-target proteins, and various repositories have cataloged these interactions from different angles. We used 4 databases to identify genes interacting with mental health drug chemical compounds (which we refer to as DrugGenes): the Drug Repurposing Hub (DRH; 3/24/2020 version),[95] the Drug-Gene Interaction Database (DGIdb; 12/2023 version),[70] the BioGRID database of protein-chemical interactions (v4.4.236),[35,37] and DrugCentral (Postgres v14.5; 11/01/2023).[67] For the DGIdb, drug-gene pairs with low interaction scores (<0.50) were filtered to align with other studies’ QC procedures.[31,96,97] As we linked these drugs into the interactome based on their drug-protein interactions, edge-weights were assigned to reflect the accumulation of evidence across databases in which they were identified (i.e., an edge that was identified in 1 database received a weight of 0.25, 2 databases corresponded to 0.5, 3 databases corresponded to 0.75, and 4 databases corresponded to 0.99 (**Supplementary Data 2**).

### Prioritization of Network Communities

Identifying communities with strong network homophily or direct paths to the mental health drugs was another central goal for our analyses. For measuring network homophily, we used the same cosine similarity score matrix that was previously described. We averaged the cosine similarity between a given vertex and a drug’s set of DrugGenes (using the RMS) and then averaged those scores (using the RMS) across the genes in each community. We computed the cosine similarity with drugs’ DrugGenes rather than to the drug vertices themselves because the vertices were of the same category (gene products) and being on the same playing field in the interactome yielded more meaningful measures of homophily. In order to assess distance between the communities and the mental health drugs, we constructed an adjacency matrix which included the interactome and the edges between mental health drugs and their DrugGenes. We then computed the shortest distance score using the same methods that were described in the previous section: reversing the weights, using Dijkstra’s algorithm, and converting the summed path lengths into a distance score ranging from 0 to 1. We once again used the RMS to average the distance scores across vertices in each community to identify communities with direct paths to the mental health drugs. These two measures provided us with a way to evaluate each community’s network-based similarity and proximity to the mental health drugs. We now had a framework for prioritizing certain communities in the network that were pertinent to our research question: 1) a community containing genes relevant to adiposity has biological relevance to the adverse side effects of interest; 2) a community with genes that have high cosine similarity scores has evidence for network homophily (common neighbors) with the drugs’ DrugGenes; and 3) a community with genes that have high shortest distance scores has evidence for direct paths to the drugs (a potential mechanistic link).

### Connectivity Significance to Mental Health Drugs in the Adiposity Subnetwork

While the previously described methods helped us identify overall communities that were similar and proximal to mental health drugs, we were also interested in implementing another technique to identify a drug-affiliated module of genes. GRIN was a network-based method that refined a set of seed vertices to a smaller set of GRIN-genes (i.e., internal prioritization of seed vertices); in contrast, DIAMOnD[50] is a network-based method that iteratively expands beyond a set of seed vertices to identify a broader set of DIAMOnD-genes (i.e., external prioritization outside of seed vertices). The DIAMOnD algorithm is fundamentally based on a statistical measure of connectivity significance for nodes linking to a module that grows with each iteration. In the first iteration, the non-seed vertex with the highest-ranking connectivity significance to the seed-based module is added, and each iteration thereafter repeats the same greedy process. The resulting set of DIAMOnD-genes provides a rank ordering of vertices with corresponding p-values. These connectivity p-values come from the cumulative probability of the hypergeometric distribution (comparing the number of observed edges between a candidate node and the seed-based module versus what would be expected by chance given the node’s degree). We implemented DIAMOnD using an unweighted version of the adiposity subnetwork with links to the mental health drugs and default parameter value for the weight of the seeds: α = 1. This subnetwork was comprised of 33 communities with evidence for enrichment of adiposity-associated genes – 27 communities with at least suggestive enrichment (p < 0.01) among adiposity gene sets and 6 other communities with a monogenic obesity gene. There were 2914 gene product vertices with 33,372 edges, 59 drug vertices (33 antidepressants, 26 antipsychotics) connecting to 280 DrugGenes through 1699 drug-protein edges (**Supplementary Data 3**). The 59 drug vertices were specified as the initial set of seed nodes for the algorithm’s starting point, and the other 2914 nodes were rank ordered by each successive iteration of the algorithm. The top 329 vertices that were added to the growing module had significant p-values (p < 0.05), and the 330^th^ iteration was the first observed p-value that was insignificant (**Fig. S8**). The resulting set of 329 DIAMOnD-genes were investigated to provide further context for how genes in adiposity-enriched communities connect to the antidepressant and antipsychotic drug vertices. We annotated these DIAMOnD-genes based on F2, F4, or monogenic trait associations, community membership, and used NDEx IQuery[51] to characterize their biological function.

### Random Walks with Restart (RWR)

Within our network there were 1574 genes that were associated with F2, F4, or monogenic obesity (**Tables S4-5**). We were interested in further exploring the vertices that lay on the paths between these trait-associated genes and the mental health drugs. In other words, we were interested in extending our previous analyses to identify vertices and edges with high betweenness. One could imagine all of the F2, F4, or monogenic nodes lined up vertically on the left side of the network and all of the drug nodes lined up vertically on the right side of the network. The natural question that follows this arrangement is: what are the most commonly used paths going through the network that link these two entities? To answer this question, we applied a random walk with restart (RWR) analysis. Each random walk was initialized at a randomly chosen F2, F4, or monogenic vertex, and at every step the next vertex was chosen probabilistically according to the normalized edge weights emanating from the current vertex. The random walk was prevented from traversing the same node twice (i.e., loops) by setting that edge probability to 0 after it had been visited. The random walk was absorbed (i.e., completed) when it arrived at a drug vertex. In addition, we implemented a restart parameter, r = 0.5, for the probability that at any given step the random walk would restart at the initial F2, F4, or monogenic vertex. This restart feature provided an effective strategy for controlled exploration of network connectivity between adiposity-associated genes and mental health drugs[98] as the persistent likelihood of restarting prevented us from recording random walks that wandered aimlessly away from the drug vertices into distant parts of the network. Furthermore, by utilizing a random walk framework we were able to get a sense of vertices with high betweenness that weren’t necessarily always on the shortest path – potentially of interest for off-target effects of drugs. We implemented r = 0.5 because it resulted in intermediate path lengths of a reasonable size (**Fig. S10a**); r values closer to 1 were converged on the shortest path, and r values closer to 0 returned walks with meandering and lengthy paths. To prevent dead-ends, the walk was re-initiated at a different starting vertex if the random walk restarted more than 3000 times. In order to focus these random walks on edges of the interactome with strong literature-based evidence that we could subsequently interpret directionality and inhibiting/activating action, we filtered the interactome to edges in the OmniPath literature-curated human signaling interaction network.[36], comprised of 6034 vertices and 19,659 edges. We recorded 15,000 random walks with restart. Of the 645 starting vertices in the subnetwork, the most frequently randomly selected starting node was chosen 46 times while the least frequently selected staring node was chosen 4 times, with a median of 24; the distribution was roughly normally distributed.

These random walks were then used to construct a new betweenness network based on edge-traversal frequency (scaled to range 0 to 1) in order to identify edges that were funnel points for the walks – edges that were often traveled on and had many paths leading into them were potentially key steps in the biological network that effect drug wADEs. We excluded edges with very low traversal frequency (≤4) that were neither the first step of the random walk nor the last step of the random walk: we plotted edge frequency and identified a knee-point at 4 using the kneedle algorithm[83] (**Fig. S17**). Following this pruning step, we removed pendant vertices that linked only to a starting vertex or a drug vertex for cleaner visualization. Edge weights for this network were assigned based on traversal frequency and ranged from 0 to 1, presenting a traffic heatmap for the 15,000 RWRs. There were 1609 vertices and 4942 edges in this RWR network (**Fig. S11**). To focus on central vertices and edges within this RWR network we were interested in high degree nodes which served as transit hubs (many paths leading to/from those vertices) and highly weighted edges which served as major conduits (frequently traversed by the random walks). We defined a ‘traffic score’ to integrate these two concepts, where *w* denotes the edge weight (reflecting edge traversal frequency) and *k_1_* and *k_2_* represent the weighted degrees of the nodes incident to the edge: *traffic score = w*sqrt(k_1_^2^ + k_2_^2^)*. This traffic score was scaled to range from 0 to 1 (**Fig. S10b**) and was used to identify edges and nodes with high betweenness. These edges with high traffic scores were funnel points for the random walks as they traversed the network from adiposity genes toward the mental health drug vertices. To understand the biological pathways that were relevant to an edge with a high traffic score, we plotted the 2-step ego networks surrounding those edges and performed IQuery[51] gene set enrichment analyses.

## Data Availability

All data are available in the main text or the supplementary materials, and all code used in the analysis is available at: https://github.com/char4816/MHD_Interactome

## Acknowledgments

We thank the generous feedback from members of the Institute for Behavioral Genetics (IBG). Data storage for this project was supported by the PetaLibrary and computational analysis was supported by the Blanca and Alpine high performance computing resources at the University of Colorado Boulder (funded by the University of Colorado Boulder, the University of Colorado Anschutz, and Colorado State University). CHA was supported by the IBG NIMH T32MH016880 training grant, by the Interdisciplinary Quantitative Biology program, and Hevolution/AFAR HEV∼NI23013. LME was supported by R01AG046938 and Hevolution/AFAR HEV∼NI23013. We gratefully acknowledge the various databases that were used (STRING, In Web, BioGRID, HIPPIE, OmniPath, GNCEM, Harmonizome, MEDI-C, OnSIDES, DrugBank, DGIdb, DRH, BioGRID, DrugCentral, and NDEx iQuerry) for making their data publicly available.

## Funding

IBG NIMH T32MH016880 training grant (CHA)

Interdisciplinary Quantitative Biology program (CHA)

National Institutes of Health grant R01AG046938 (LME)

Hevolution/AFAR HEV∼NI23013 (LME & CHA)

## Author Contributions

CHA and LME contributed to conceptualization, methodology design, investigation, visualization, and writing (original draft, review, and editing). CHA led the analyses and drafting of the manuscript, and LME supervised the study.

## Supplementary Materials

Supplementary information is available at MP’s website: Fig. S1-19, Table S1-S16, and Supplementary Data 1-3 (Excel files).

