## Supplementary Information for "Biological network identifies pathways linking mental health drugs to adverse weight-related side effects"

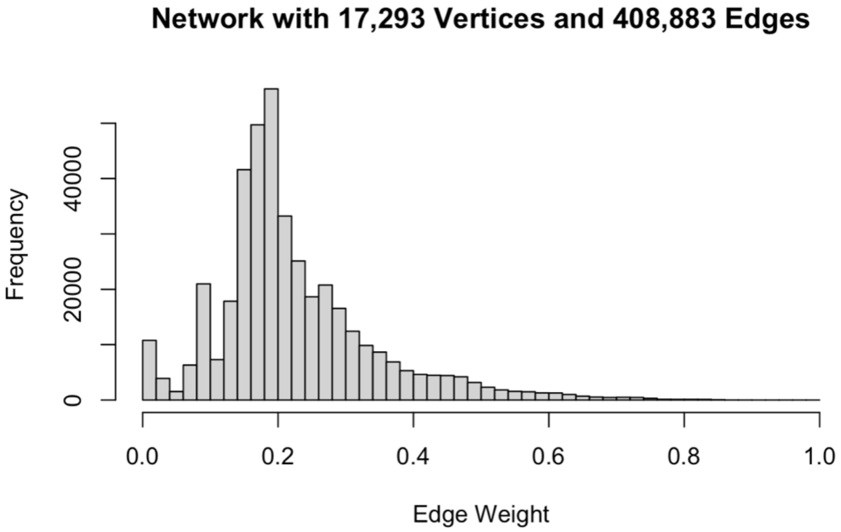


**Fig S1: Distribution of interactome edge weights.** Larger edge weights reflect increased evidence across within and between these databases (e.g., high-throughput protein interaction assays, curated pathways, text mining, co-expression, and interlogs).

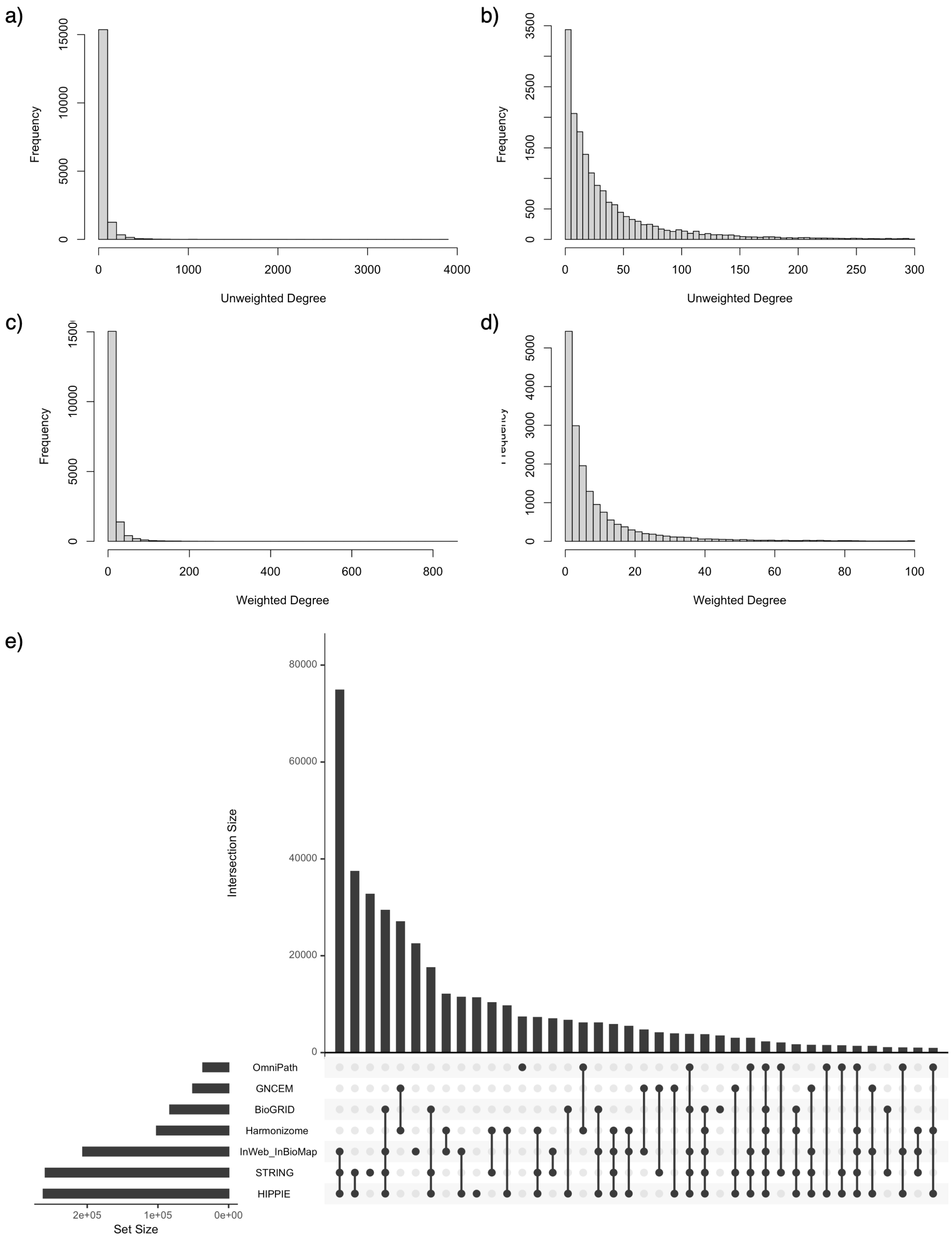


**Fig S2: Interactome degree distributions and database overlap.** Panel a depicts the unweighted degree distribution across 17,293 vertices (408,883 edges), with panel b zooming in on the x-axis (0 to 300). Panel c depicts the weighted degree distribution for the interactome with panel d zooming in on the x-axis (0 to 100). Panel e shows an upset plot depicting the overlap among databases that were used to build the interactome. 81% of the edges were identified by at least 2 of the 7 databases.

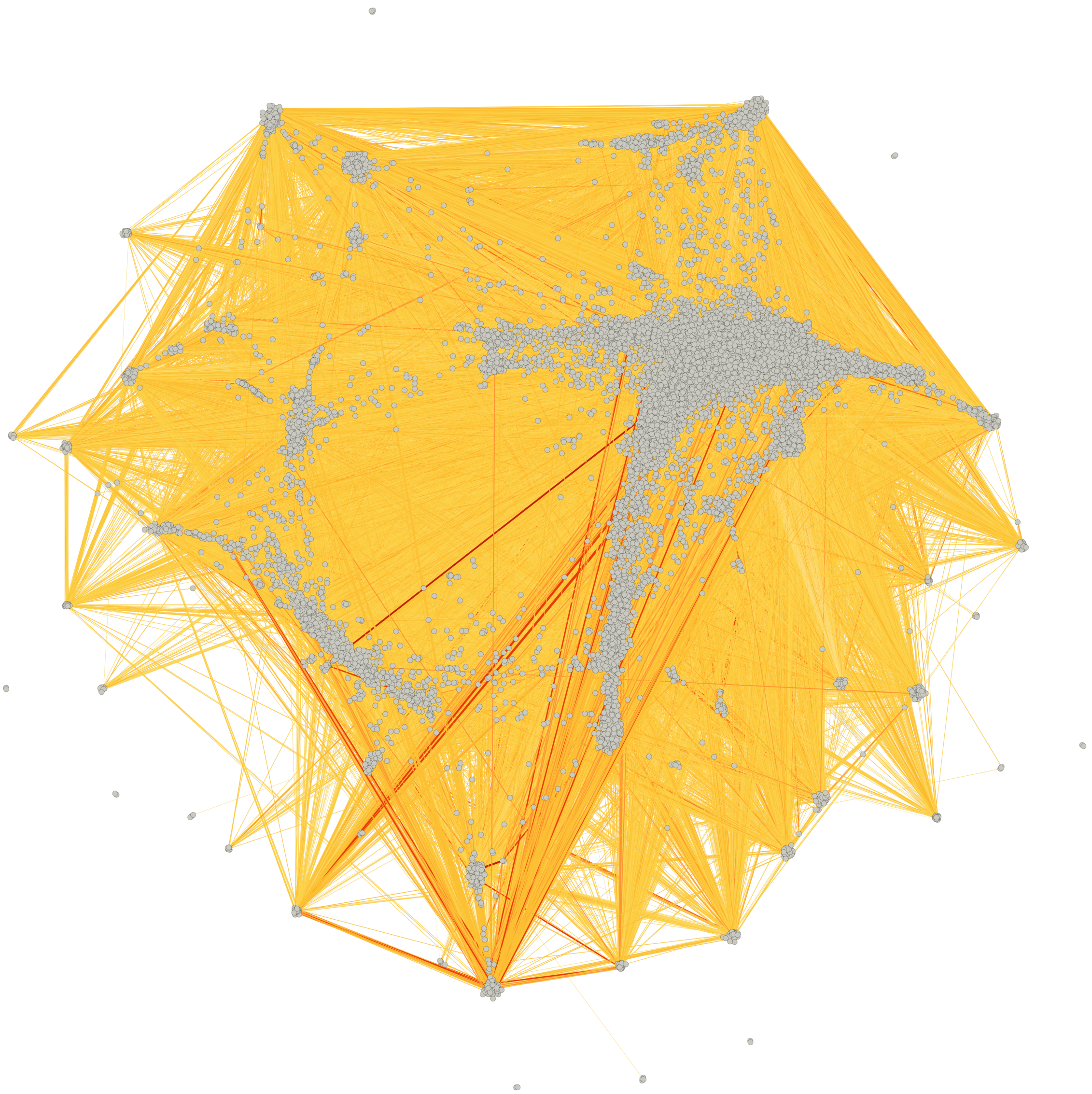

**Fig S3: Improved biological interactome.** The biological interactome consisting of 17,293 proteins/genes (vertices) and 408,883 interactions (edges). Edges with stronger interaction weights are darker red. The dense layout results in many edges being obscured beneath overlapping connections.

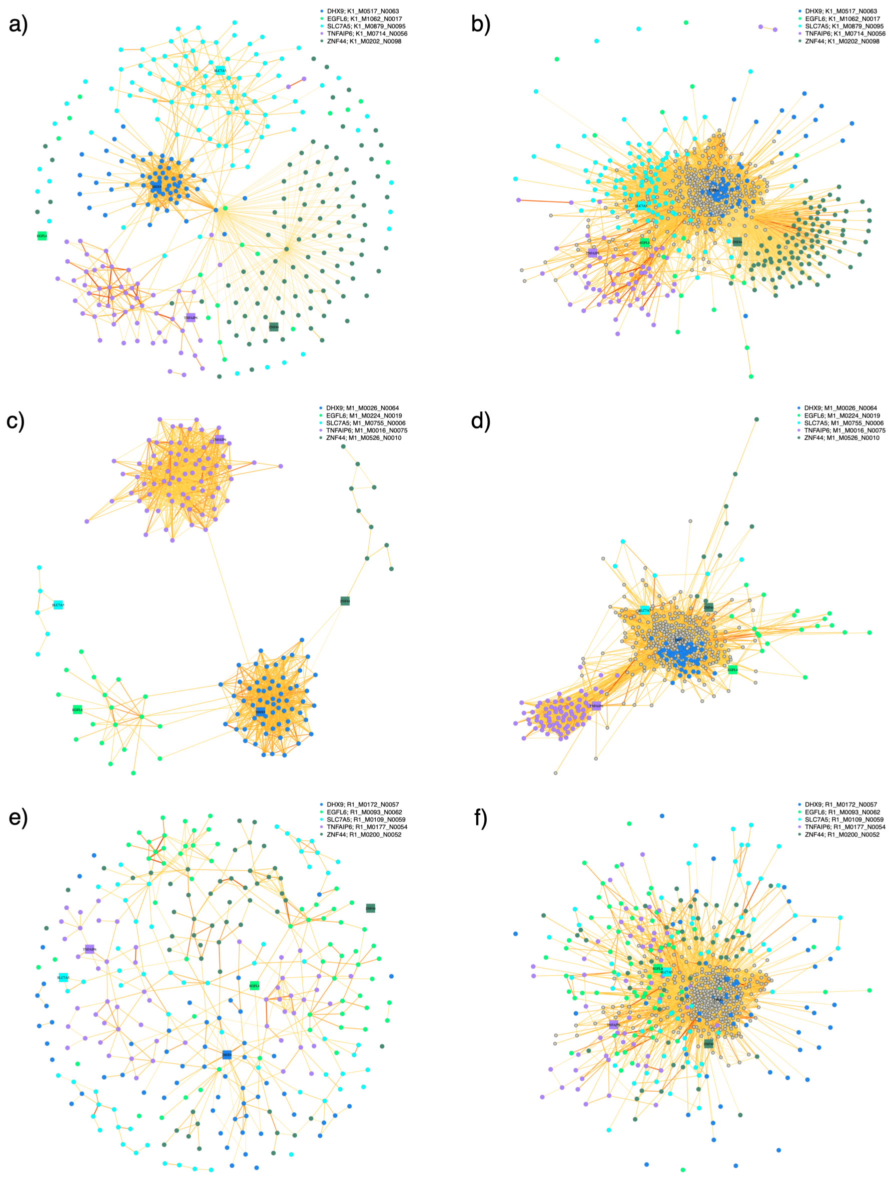


**Fig S4: Comparing module detection algorithms.** Module assignments for 5 randomly selected vertices are shown for the 3 MONET clustering methods: K1 (panels a and b), M1 (panels c and d), and R1 (panels e and f). Panels a, c, and e show the modules containing the 5 vertices in isolation (DHX9 module in blue, EGFL6 module in green, SLC7A5 module in cyan, TNFAIP6 module in purple, and ZNF44 module in green) while panels b, d, and f show those same modules with links to the surrounding vertices (colored grey) that neighbor any of those 5. These visualizations were performed for many sets of 5 randomly selected vertices, and the M1 method consistently produced the best modules (as seen in panel c) – the K1 method often included vertices with no connections to the rest of the module (panel a), and the R1 method generally defined modules that did not appear topologically distinct (panel e).

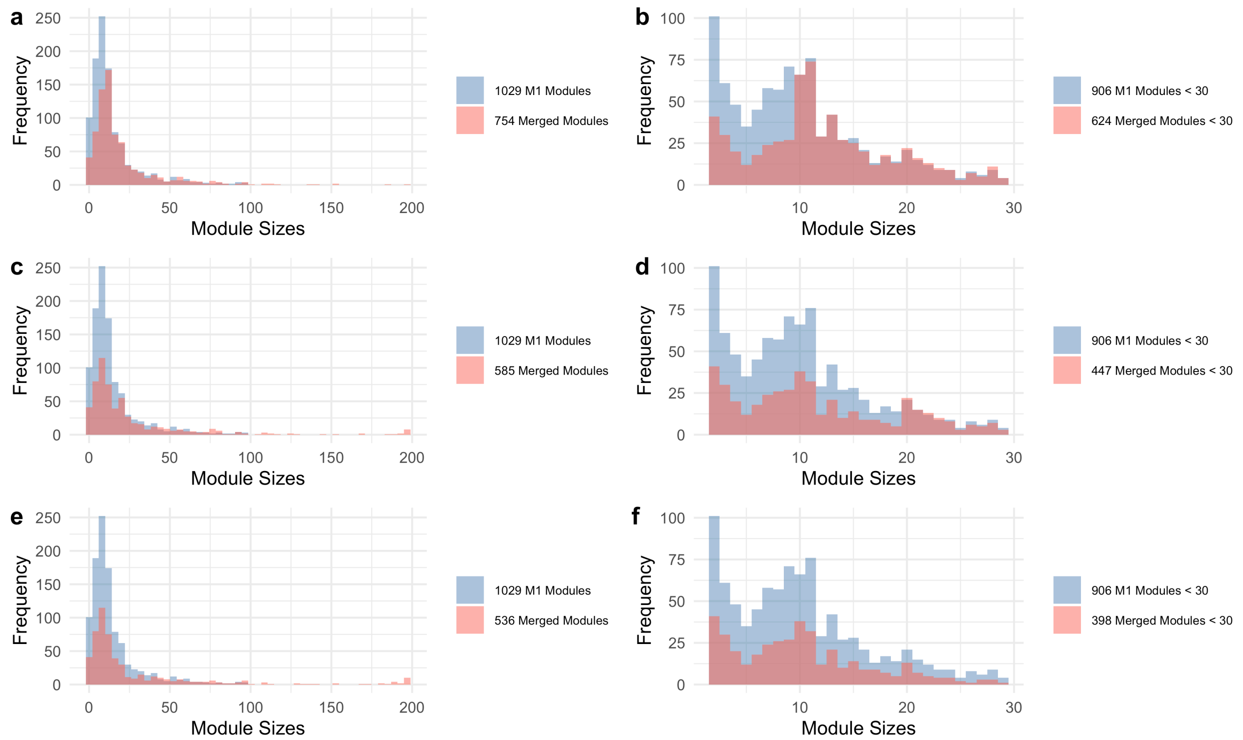


**Fig S5: Merging modules in 3 phases.** The 6,743 pairs of modules with high similarity scores and distance scores were rank-ordered by a combined Euclidian score, sqrt(similarity$^2$ + distance$^2$), and assessed for potential merging in 3 phases based on community size. First, pairs of modules were considered for a merge if either of the modules was sized < 10, then we considered merging modules if either was sized < 20, and finally we considered merging modules if either was sized < 30. In each of the 3 stages all 6,743 pairs of modules were considered in rank order, and merges were not allowed if the sum of both module sizes was ≥ 200. Histograms of module sizes are shown for each of the 3 merging phases (panels a-b for phase 1, panels c-d for phase 2, and panels e-f for phase 3) with the original M1 module sizes in blue and the post-merging community sizes in red. The histograms on the right (panels b, d, and f) zoom-in on the smaller modules (size < 30). Our method for combining small modules with their similar neighboring modules resulted in 536 communities which we used for downstream analyses (e.g., enrichment tests).

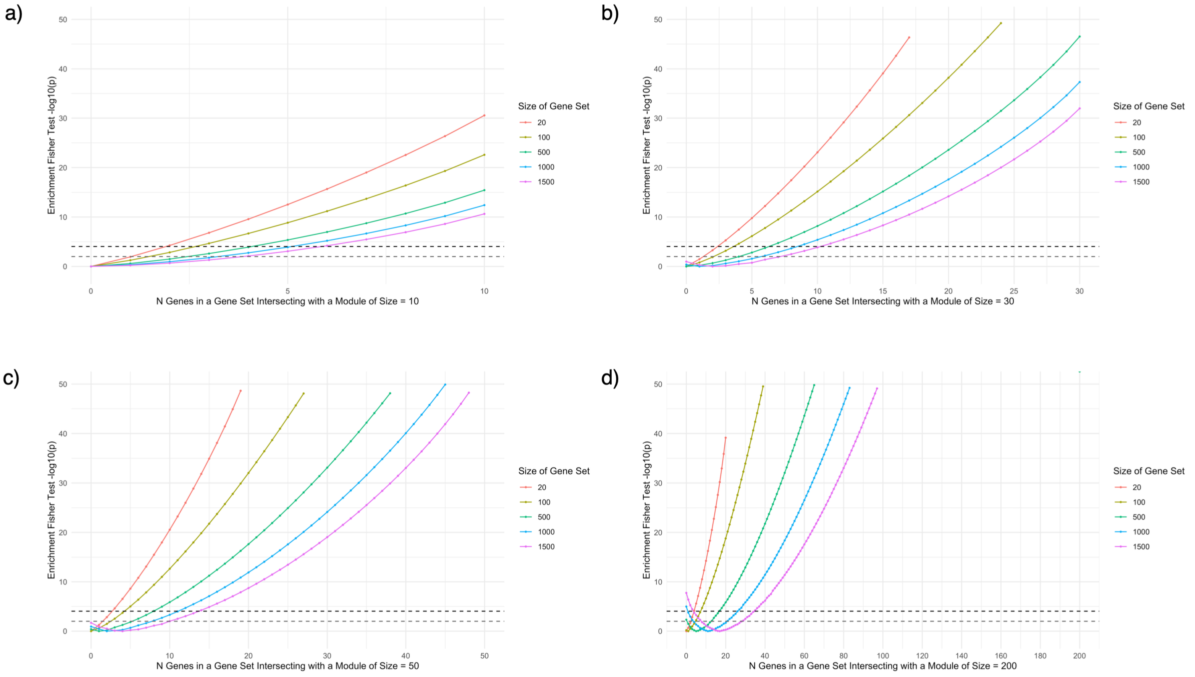


**Fig S6: Simulated enrichment tests for modules.** Simulations for Fisher's exact tests to assess enrichment of adiposity gene sets in a module with 10 vertices (panel a), 30 vertices (panel b), 50 vertices (panel c), and 200 vertices (panel d). Modules with less than 10 vertices showed little ability for significant results at the Bonferroni-corrected significance threshold ($p < 9.33 \times 10^{-5}$, black dashed line) and suggestive enrichment threshold ($p < 0.01$, grey dashed line), especially for larger gene sets. Modules with at least 20-30 vertices were more reasonably sized for enrichment tests.

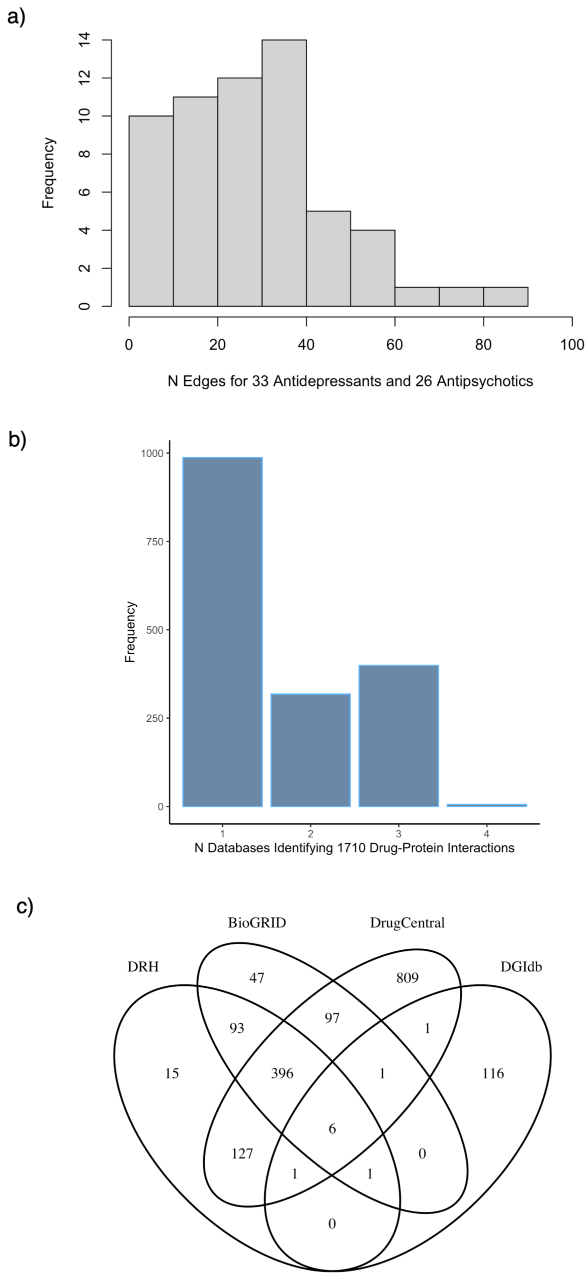


**Fig S7: Drug Links to the Interactome.** We evaluated the links between our interactome and 59 mental health drugs (33 antidepressant and 26 antipsychotics). Panel a illustrates how some drugs were highly promiscuous with more than 60 protein interactions (e.g., chlorpromazine, haloperidol, and olanzapine), and others were more specific, interacting with less than 5 drug targets (e.g., isocarboxazid, pimavanserin, desvenlafaxine, and moclobemide). The majority of these mental health drugs (38 out of 59) had between 10 and 40 interactions. Panel b shows the redundancy of edge identification across the 4 databases (1710 edges between the interactome and these 59 drugs were found across the DRH, BioGRID, DrugCentral, and DGIdb) with more specific database overlap shown in the diagram in panel c. A notable number of edges were identified by only 1 database highlighting the importance of integrating multiple sources of information.

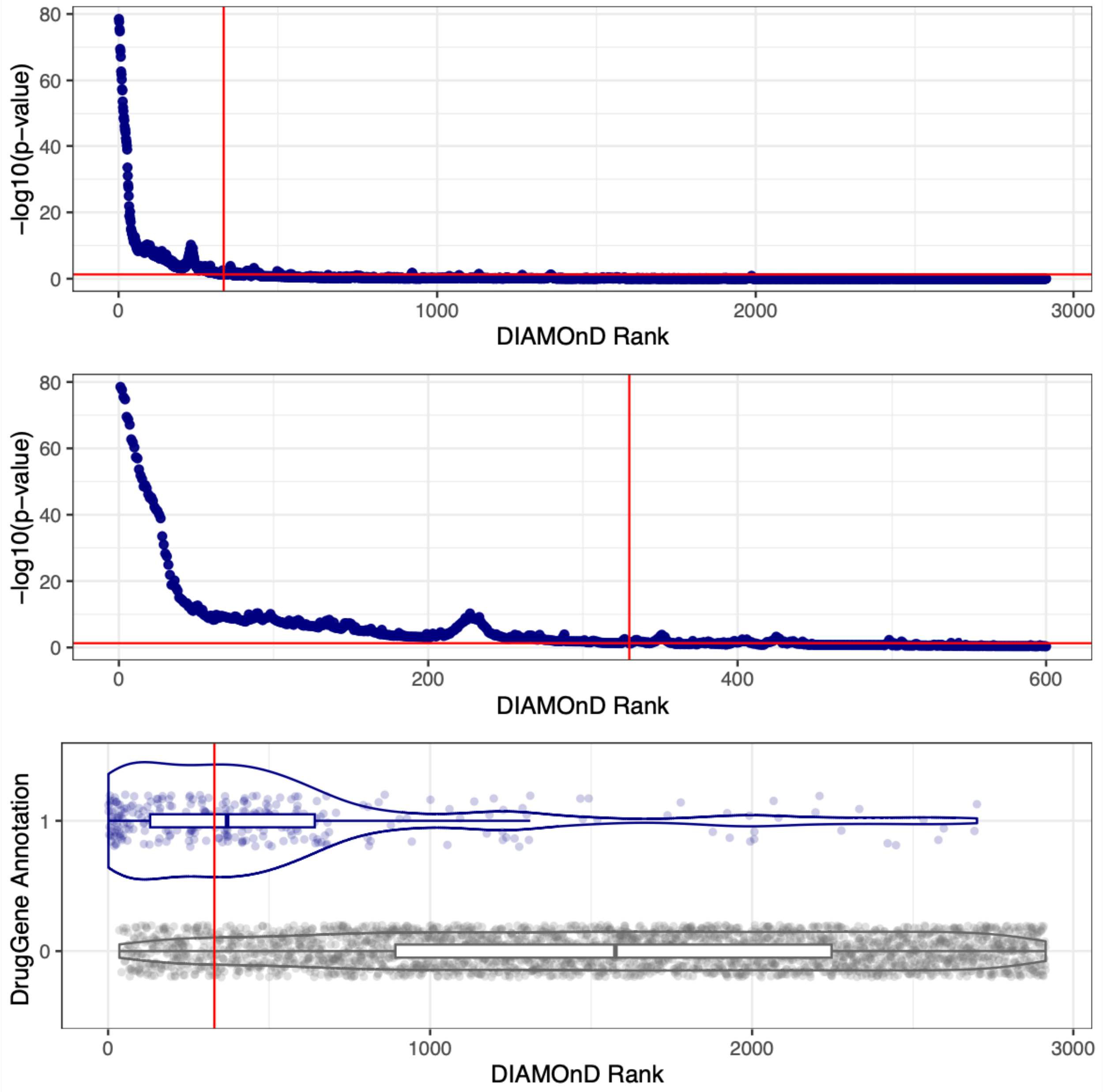


**Fig S8: Identification of DIAMOnD-genes.** The top panel shows the connectivity significance values for each iteration of DIAMOnD (on the -log10 scale), with the middle panel zooming in on the first 600 iterations. The vertical and horizontal red lines mark the threshold at which the connectivity significance p-value was less than 0.05 through the first 329 iterations. The bottom panel shows how DrugGenes (1 on the y-axis) were primarily identified by DIAMOnD in the earlier iterations.

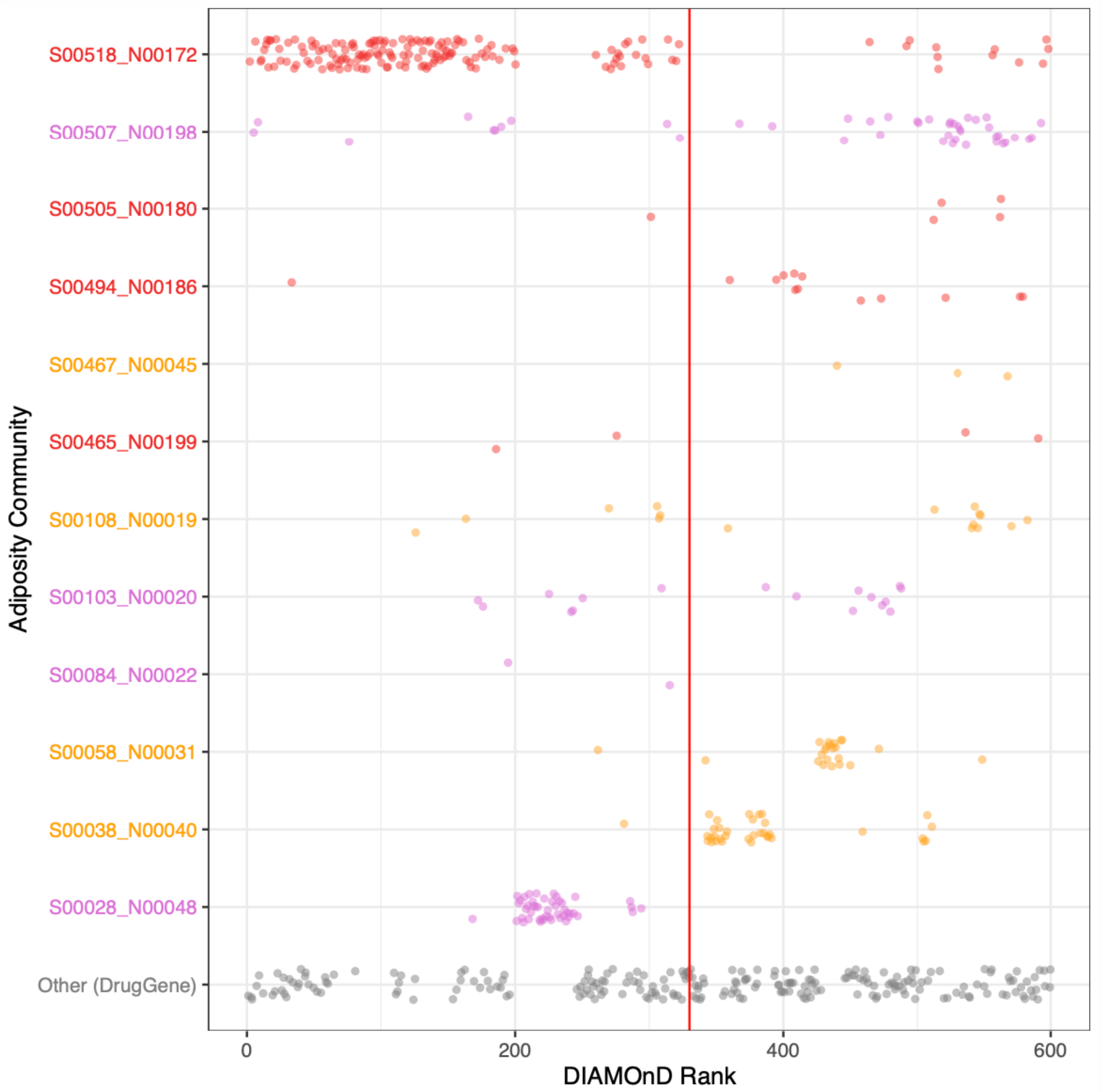


**Fig S9: Community membership of DIAMOnD-genes.** The vertices identified by DIAMOnD primarily belonged to 6 communities: S518_N172 (151/172 vertices), S028_N048 (48/48 vertices), S507_N198 (10/198 vertices), S103_N020 (7/20 vertices), S108_N019 (6/19 vertices). The vertical red line marks the threshold at which the connectivity significance p-value was less than 0.05 through the first 329 iterations.

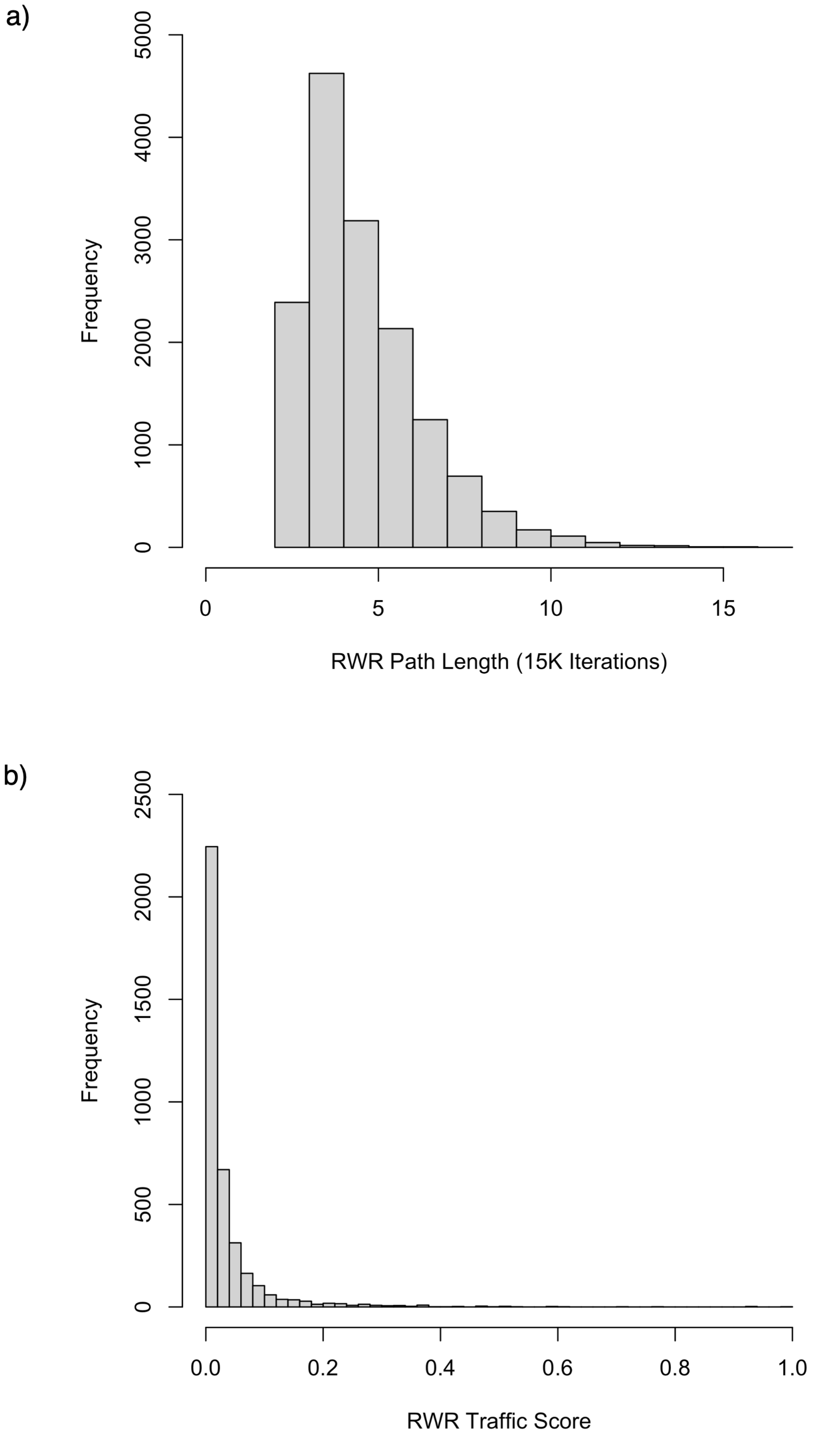


**Fig S10: RWR Distributions.** Panel a shows the distribution of unweighted path lengths for 15,000 random walks with restart. Panel b shows the traffic scores for the edges in the RWR network (mean traffic score = 0.038; interquartile range = [0.012, 0.036]).

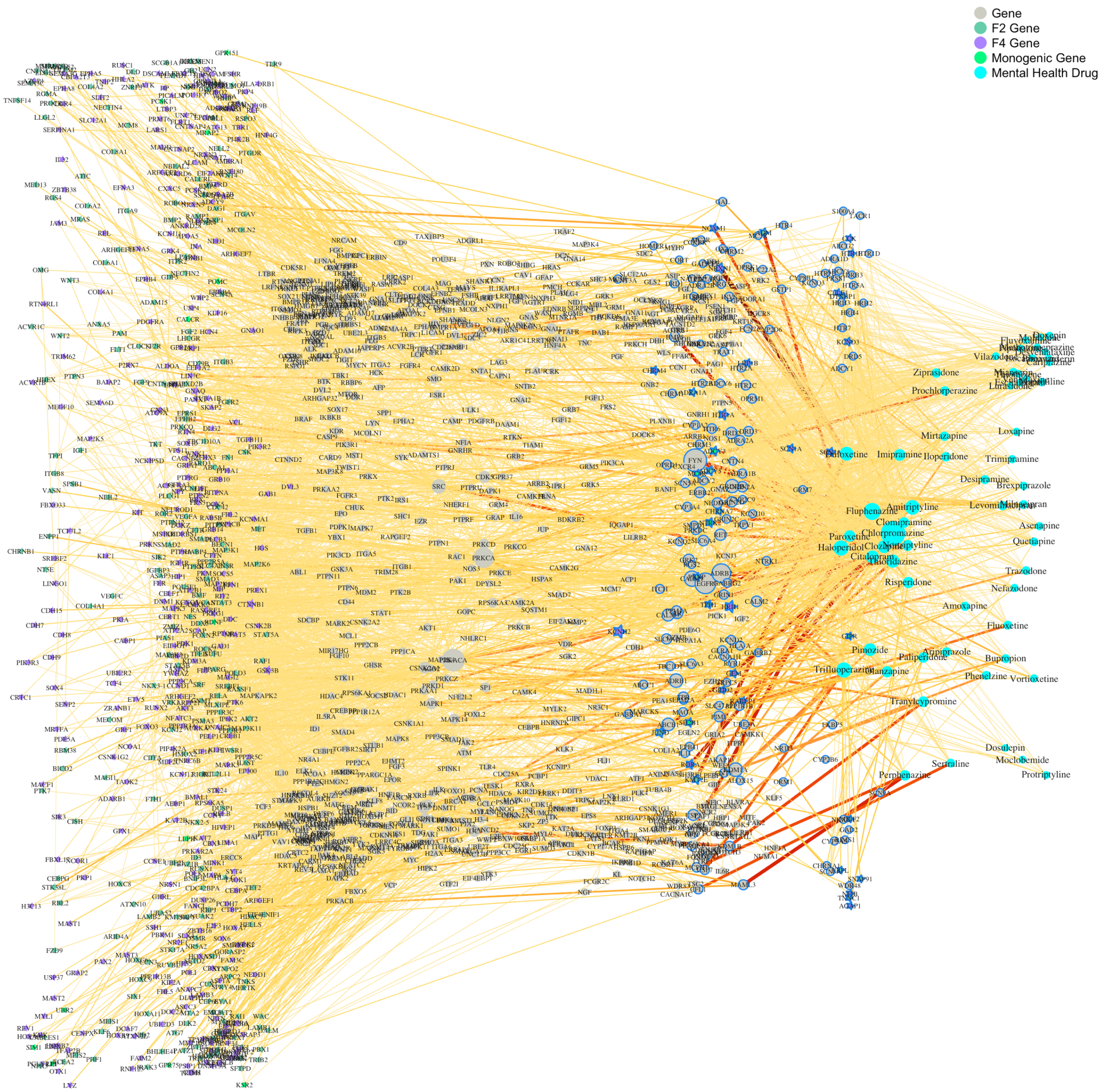


**Fig S11: Random walks with restart edge frequency.** 15,000 random walks with restart traversed the network between adiposity-associated genes and mental health drugs. The resulting network summarized the most traversed paths. Edge weights edge frequency (the proportion of random walks traversing an edge) with more frequently visited edges depicted in darker red. The larger vertices highlight bottle necks with a larger weighted degree.

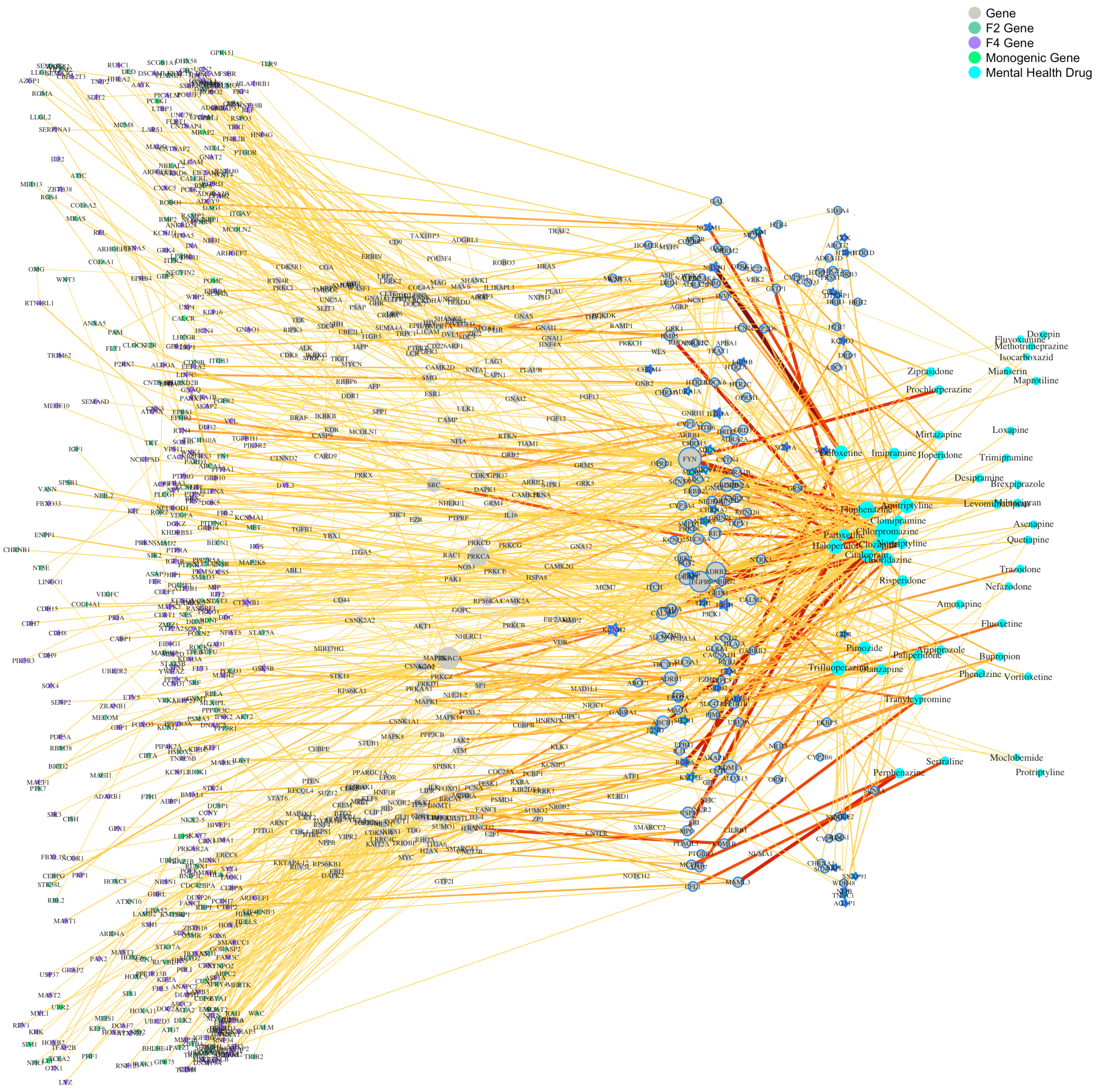


**Fig S12: Random walks with restart edge frequency, 75th percentile.** The same network as Fig SX, with edges below the 75th percentile removed. Thus, the edges and vertices that participated in many of the random walks are emphasized in this visualization.

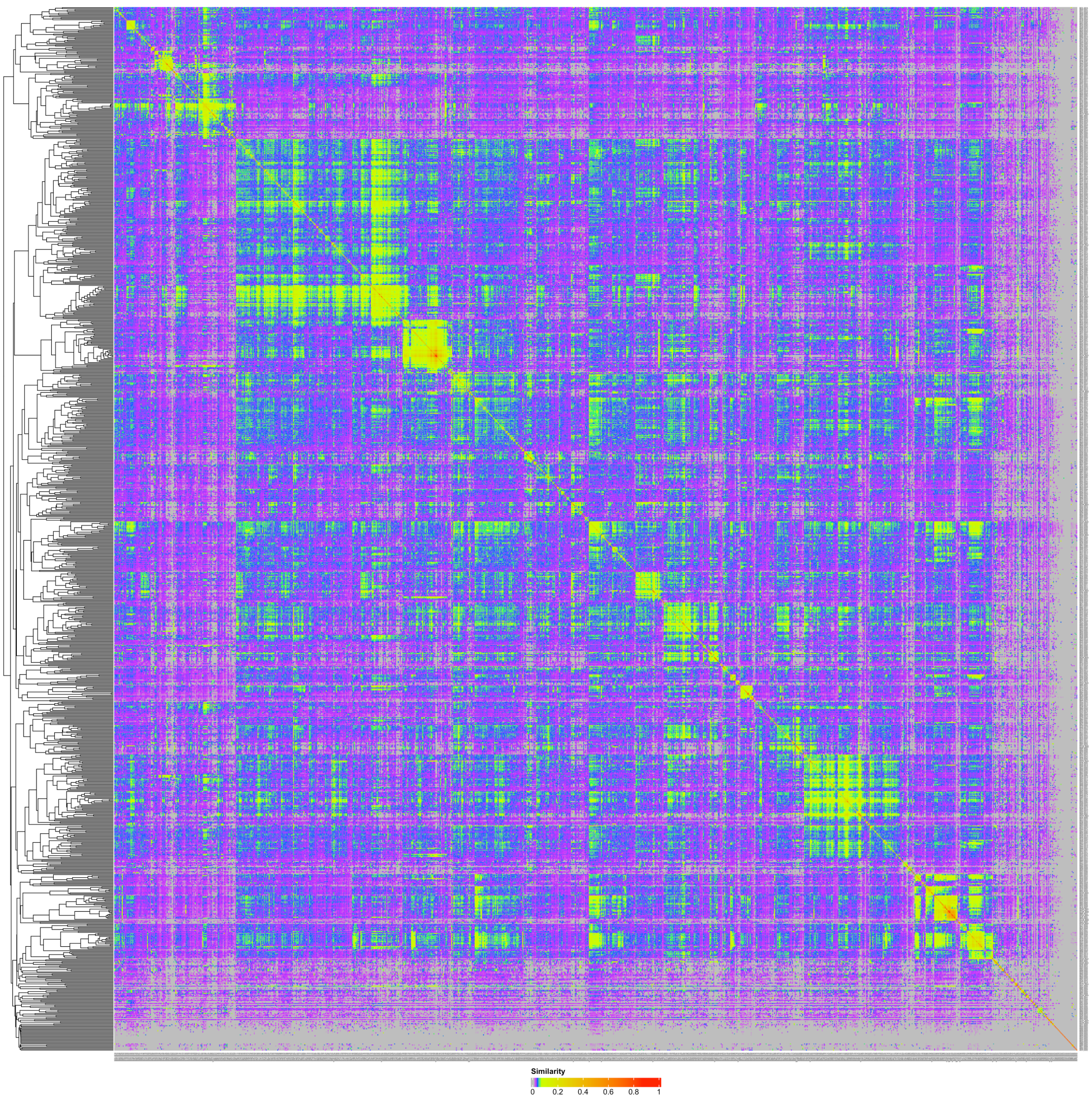


**Fig S13: Module similarity.** Heatmap showing generalized mean cosine similarity between vertices for each pairwise comparison of MONET M1 identified modules. 1029 modules are represented by the rows and columns of the symmetrical heatmap, ordered according to the dendrogram shown on the left.

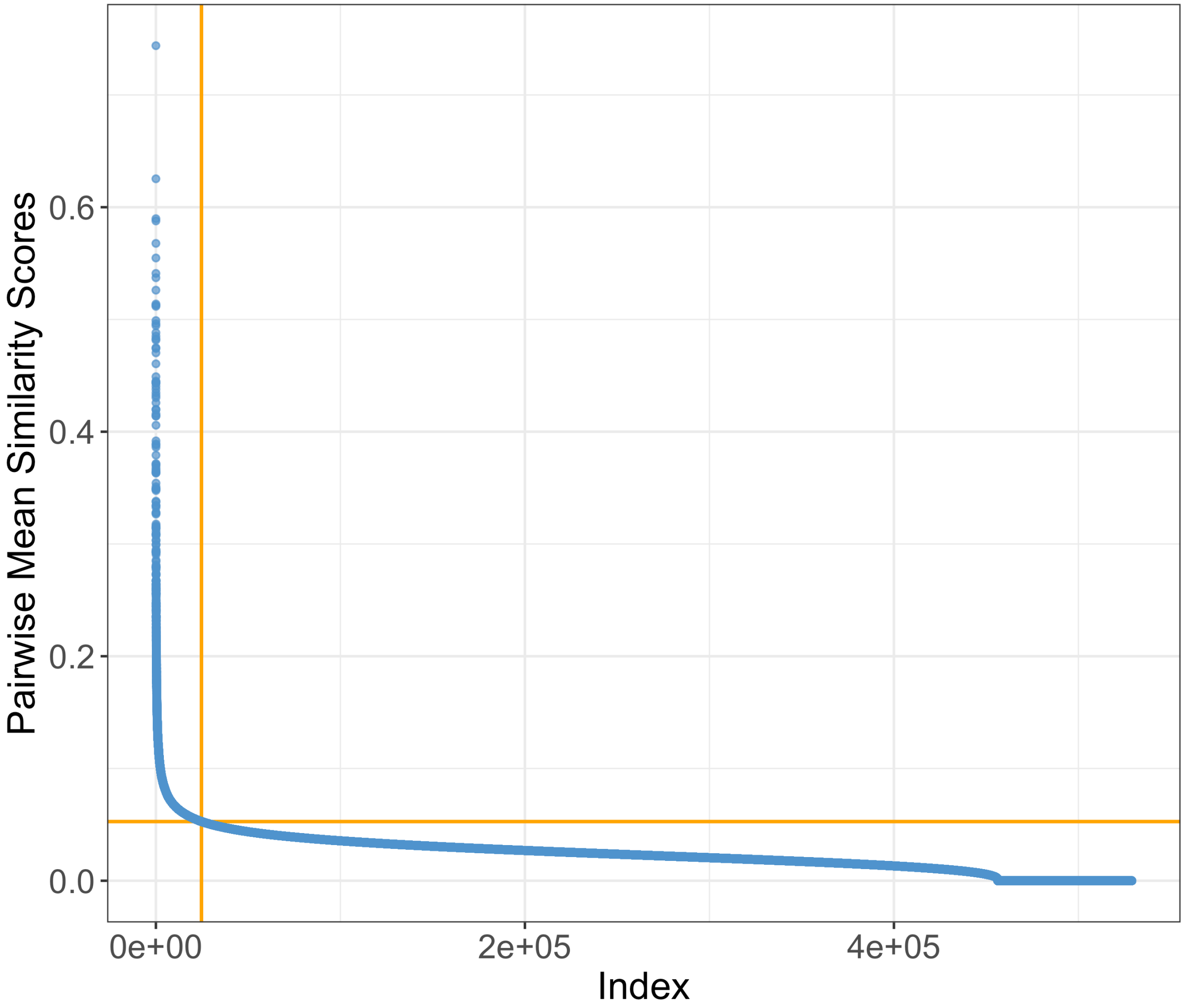


**Fig S14: Module similarity.** The rank-ordered pairwise generalized mean similarity scores between modules of vertices. Each blue dot corresponds to an entry of the lower-triangle of module similarity matrix (depicted as a heatmap in Supplementary Figure #). The kneedle knee-point identification algorithm (sensitivity = 89) was used to identify a cutoff for the 24,692 pairs of modules (left of the vertical orange line) that had a relatively larger cosine similarity (above the horizontal orange line at 0.05270676).

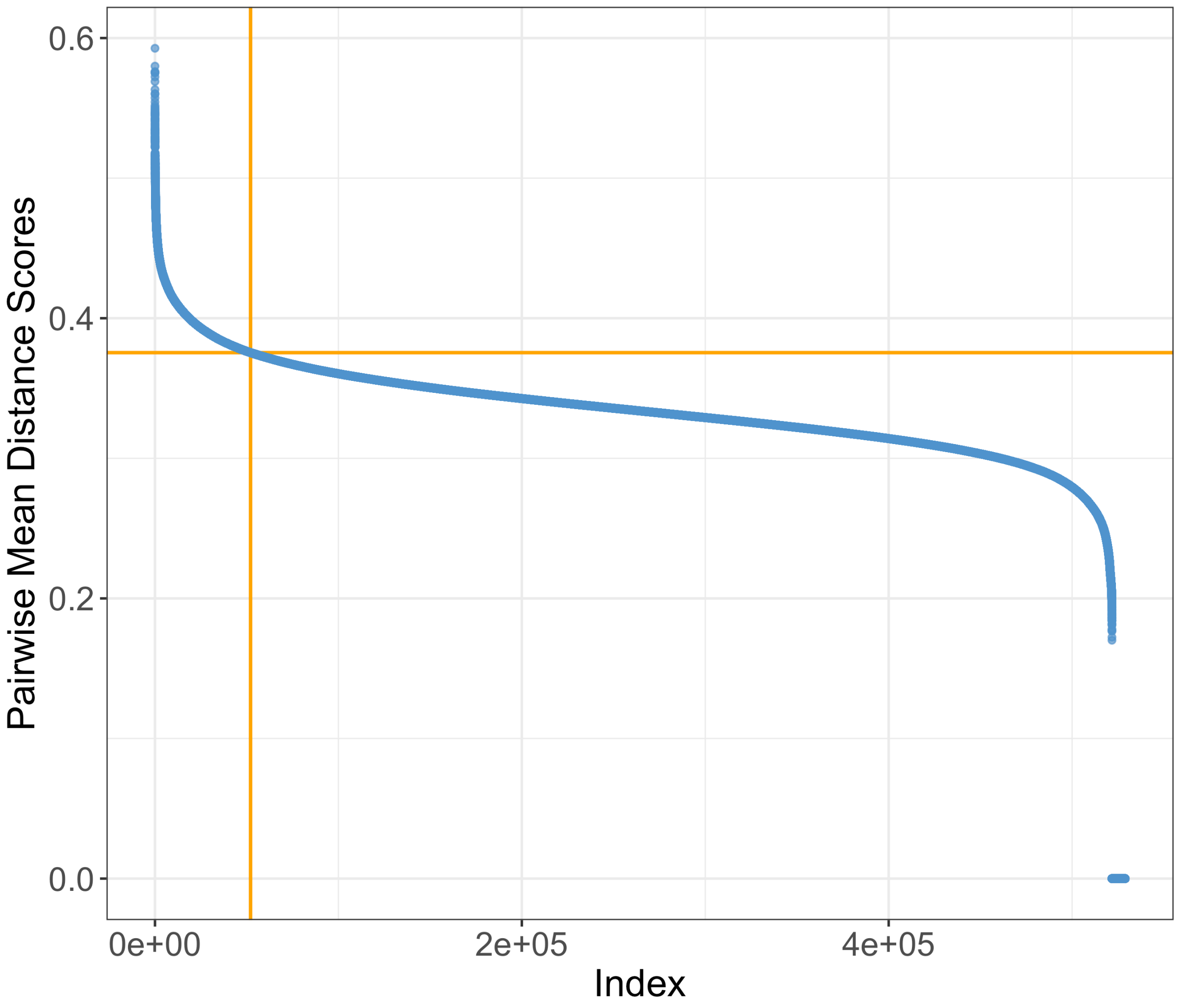


**Fig S15: Module distance threshold.** The generalized mean shortest path distance scores between vertices in 2 different modules (using Dijkstra's algorithm) are rank ordered along the x-axis. There are 528,906 blue dots indicating the generalized mean shortest distance score between vertices belonging to a pair of MONET M1 modules. The kneedle knee-point identification algorithm (sensitivity = 52) was used to identify a cutoff for the top 52,100 pairs of modules (left of the vertical orange line) that had relatively larger mean distance scores (above the horizontal orange line at 0.3754363).

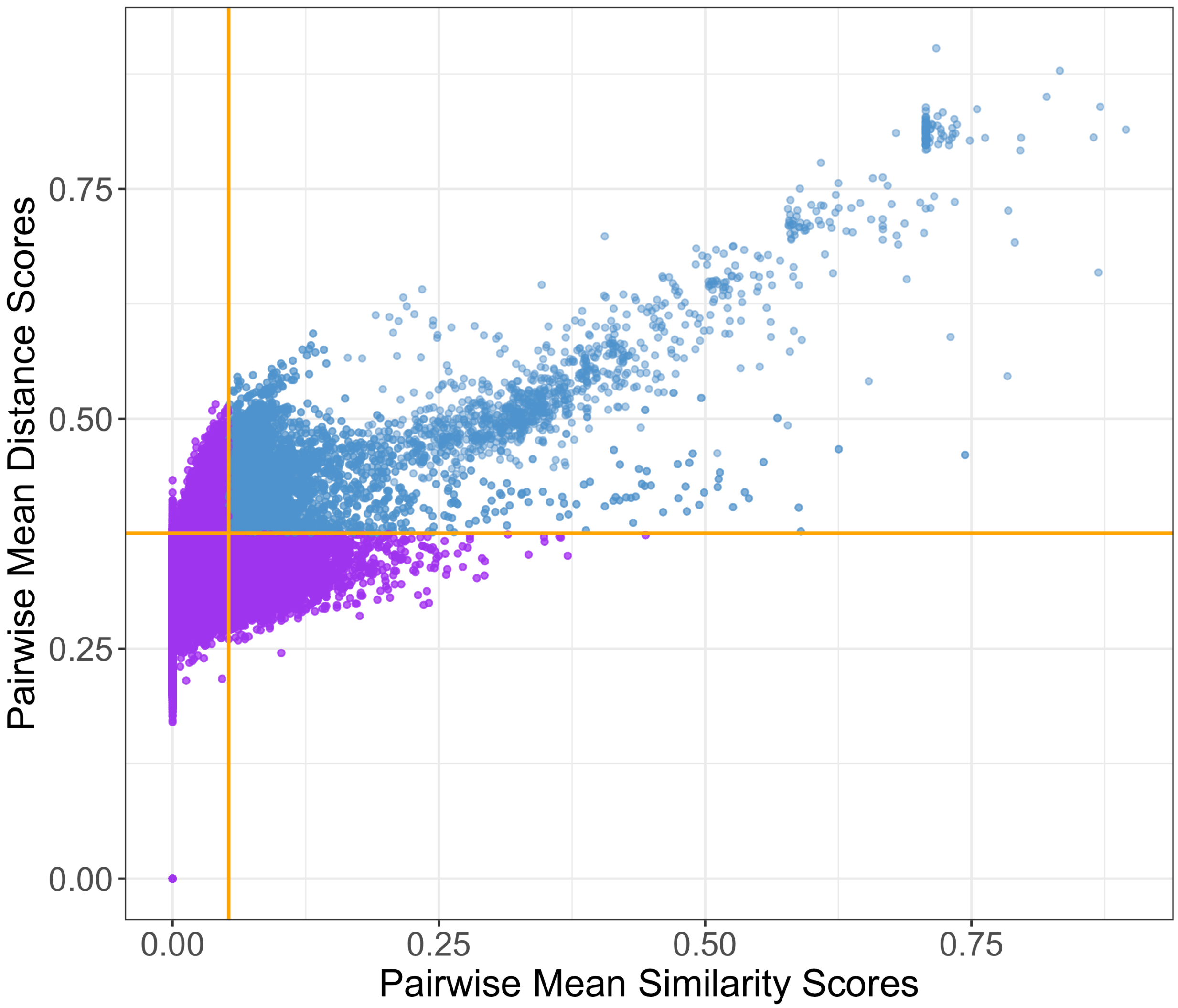


**Fig S16: Merging modules – similarity and distance criteria.** A scatter plot illustrating the similarity and shortest distance measures for each pairwise comparison of modules (each dot represents a pair of MONET M1 modules). We only considered merging a pair of modules if they exhibited high scores for mean similarity and mean distance (blue dots), and 6,743 out of 528,906 pairs of modules (1.27%) met the criteria of having mean cosine similarity ≥ 0.05270676 (vertical orange line) and mean distance score ≥ 0.3754363 (horizontal orange line). These thresholds were identified using the kneedle knee-point identification algorithm for each metric.

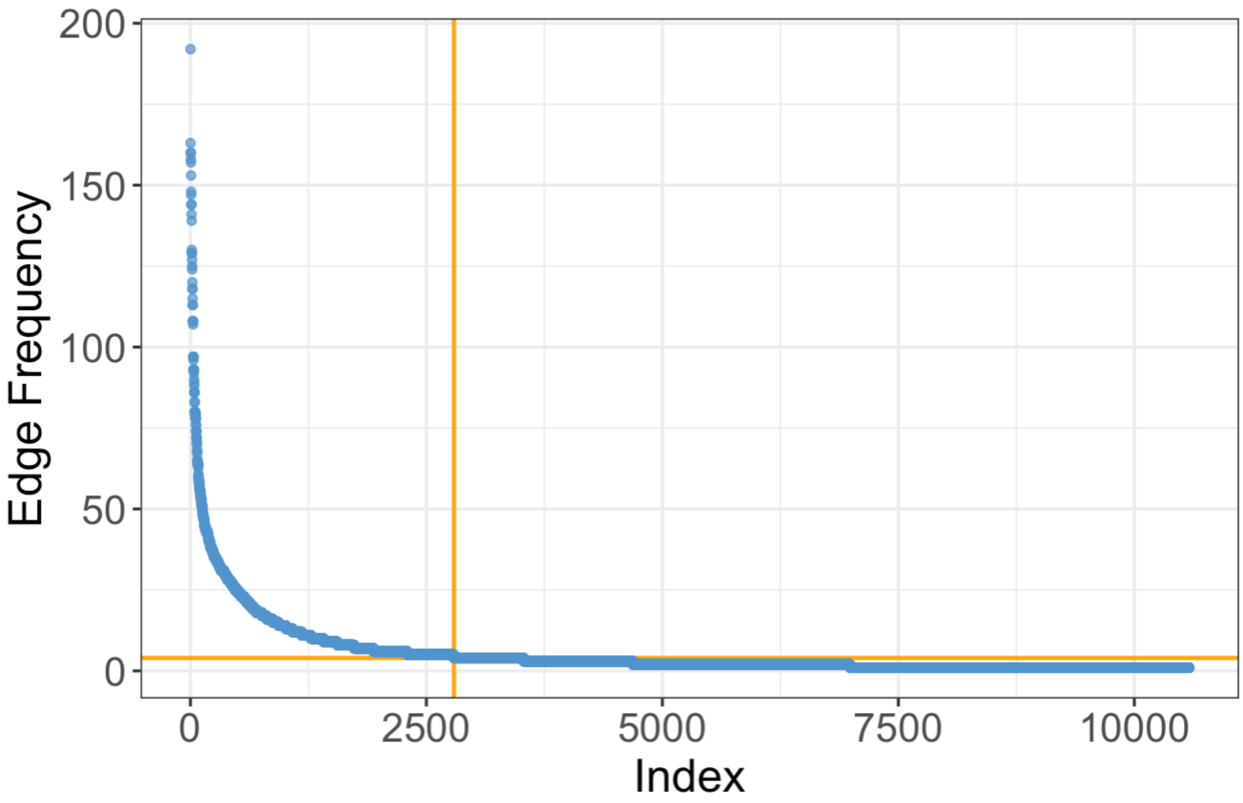


**Fig S17: Rank ordered edge frequency for 15,000 random walks with restart.** The kneedle algorithm identified an elbow point at 4 (horizontal line) for pruning the RWR network.

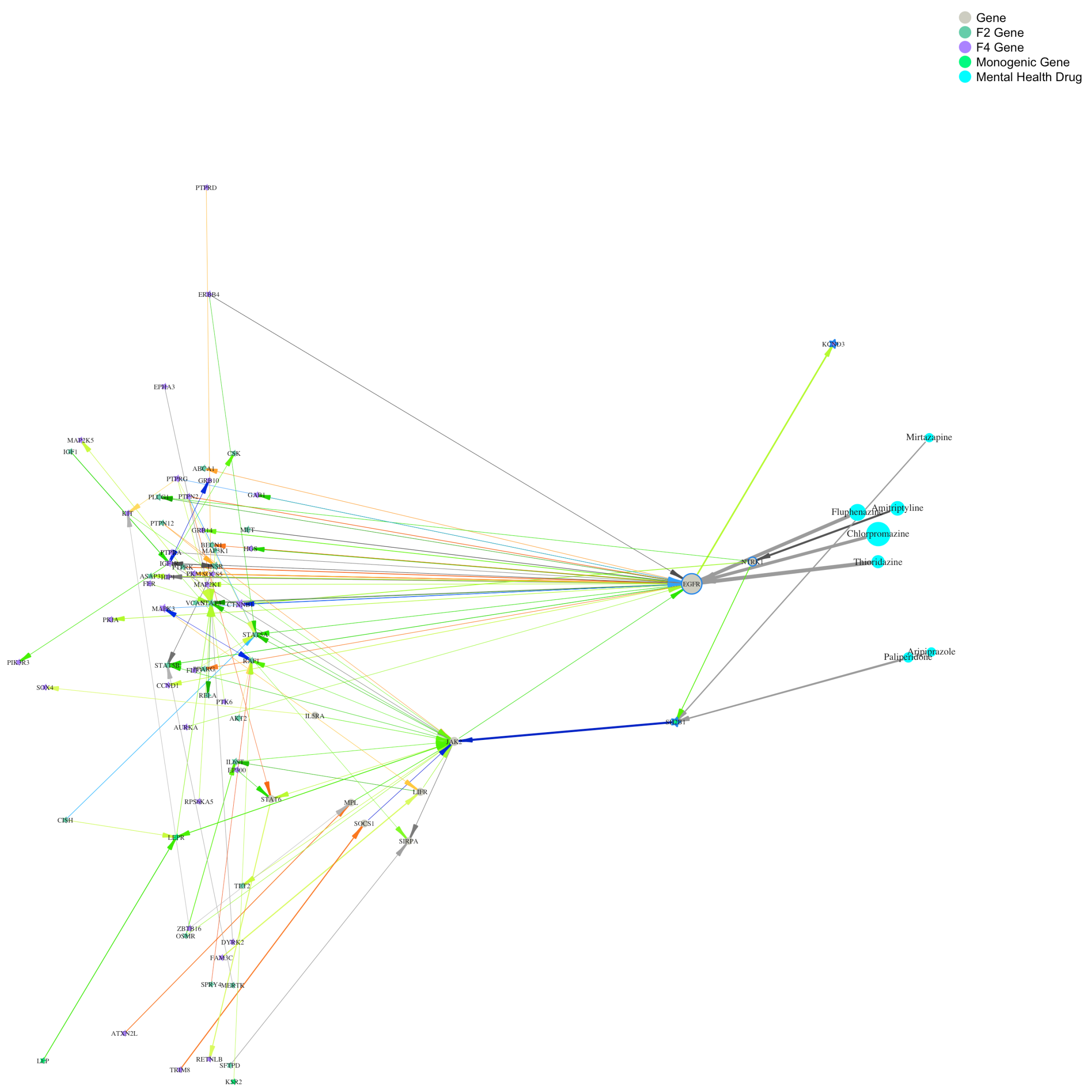


**Fig. S18: Random walks with restart highlighted the leptin signaling pathway.** The edge between JAK2-SH2B1 unveiled a 2-step ego network that was significantly enriched for the leptin signaling pathway and this plot depicts the 2-step ego network extracted from Fig. S11. Arrows indicate the directionality (or bidirectionality) from source to target vertices and colors indicate the type of action (green for stimulatory, red for inhibitory, grey for neither, and blue for both).

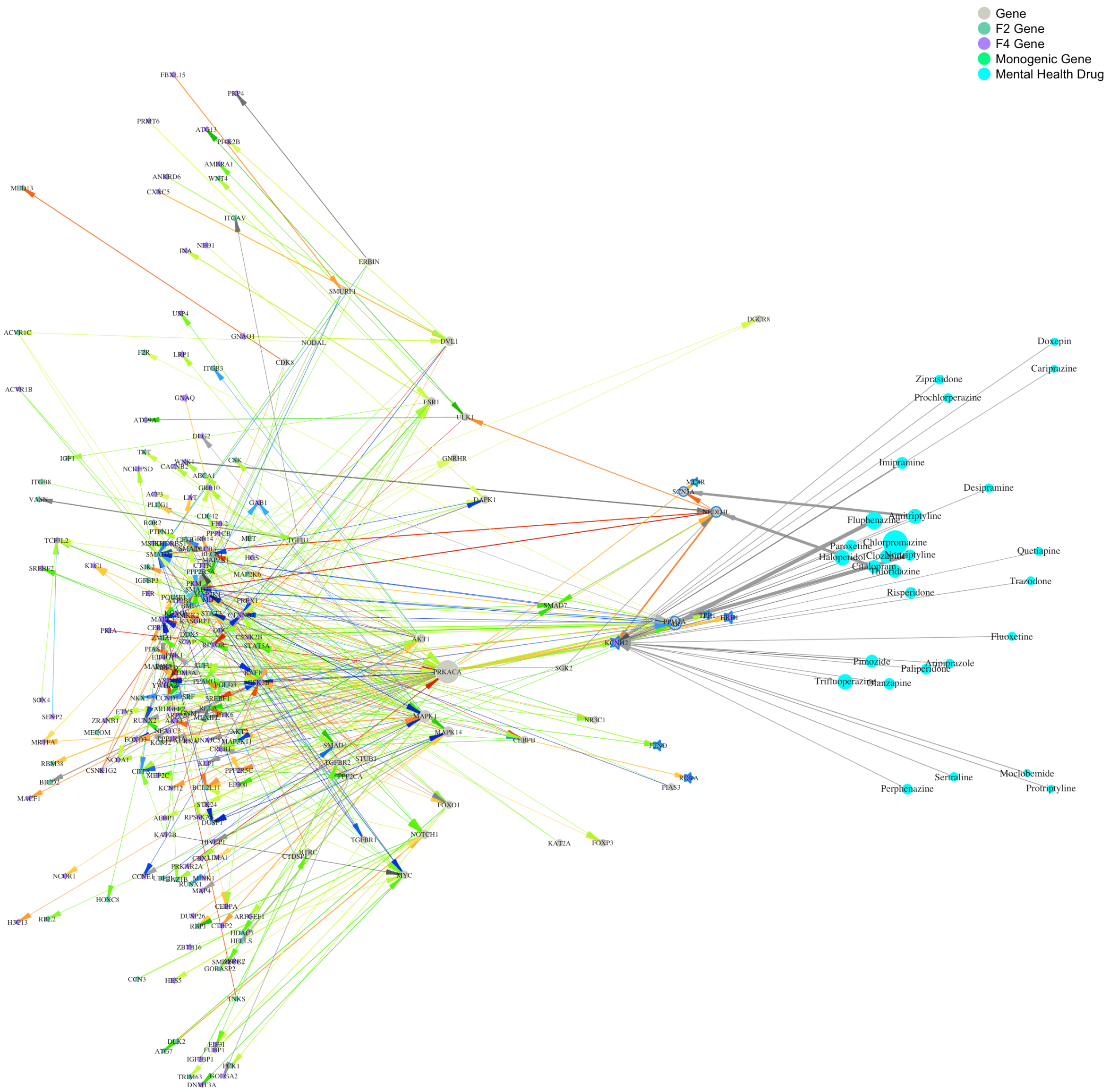


**Fig. S19: Random walks with restart highlighted the adipogenesis signaling pathway.** The edge between NEDD4L-SMAD3 unveiled a 2-step ego network that was significantly enriched for the adipogenesis pathway, and this plot depicts the 2-step ego network extracted from Fig. S12. Arrows indicate the directionality (or bidirectionality) from source to target vertices and colors indicate the type of action (green for stimulatory, red for inhibitory, grey for neither, and blue for both).

**Table S1: Databases used to construct our interactome.**

| Database | Primary or Auxiliary | Weights | Solitary Identification Threshold | Retained Edges | Considered Edges |
|---|---|---|---|---|---|
| STRING Physical*(2)* | Primary | Yes | 0.70 | 259,108 | 655,796 |
| BioGRID MV-Physical*(3, 4)* | Primary | No | -- | 82,907 | 79,035 |
| InWeb_InBioMap*(5)* | Primary | Yes | 0.112 | 206,014 | 450,083 |
| OmniPath*(6)* | Primary | No | -- | 36,156 | 36,156 |
| HIPPIE*(7)* | Primary | Yes | 0.73 | 261,937 | 802,202 |
| Harmonizome*(8)* | Auxiliary | No | -- | 101,706 | 2,508,443 |
| GNCEM*(9)* | Auxiliary | Yes | -- | 50,495 | 1,611,192 |
| | | | **Final Network** | **408,883** | **5,314,131** |

**Table S2: Databases used to construct the drug-protein interactome.** There were 4 databases that linked the 59 mental health drugs to the interactome. DrugCentral contributed the most edges linking the mental health drugs to the interactome (1438) and DGIdb contributed the least (126).

| Database | QC Threshold | Retained Edges |
|---|---|---|
| DRH *(70)* | -- | 639 |
| DGIdb *(71)* | 0.5 | 126 |
| BioGRID Chemicals *(3, 4)* | -- | 641 |
| DrugCentral *(41)* | -- | 1438 |
